# Randomised Trial of a Multilingual Conversational AI for Preoperative Education

**DOI:** 10.64898/2026.05.24.26353997

**Authors:** Yuhe Ke, Chenglin Niu, Jingchi Liao, Jacqueline Sim, Hairil Rizal Abdullah, Liyuan Jin, JingZhi An, Henry Sun Sien Ho, Joshua Yi Min Tung, Hiang Khoon Tan, Ban Leong Sng, Daniel Shu Wei Ting, Marcus Eng Hock Ong, Nan Liu

## Abstract

**Background:** Informed consent depends on patients’ understanding of anaesthesia risk, yet comprehension remains poor despite routine preoperative consultation. Conversational artificial intelligence (AI) could establish patient-reported understanding before clinician contact, but whether such systems can achieve patient-reported understanding comparable to clinician-delivered education remains unknown.

**Methods:** We conducted a randomised equivalence trial (n = 130) of PEAR (Preoperative Education of Anaesthesia Risks), a multilingual retrieval-augmented conversational AI grounded in institutional consent materials, versus standard preoperative consultation in adults undergoing elective surgery.

**Results:** A total of 130 adults (mean age 52.4 ± 14.5 years) were enrolled. Post-consultation understanding scores in the PEAR group met the pre-specified equivalence criterion compared with standard consultation across all three primary measures. Patients who interacted with PEAR before clinician contact achieved understanding scores comparable to those receiving standard face-to-face consultation alone. PEAR reduced documentation and consultation time, corresponding to a projected annual net benefit of approximately SGD 0.99 million (USD 0.78 million) at a single tertiary centre.

**Conclusions:** A retrieval-augmented conversational AI achieved patient-reported understanding of anaesthesia risk equivalent to standard preoperative consultation while substantially improving workflow efficiency. These findings support supervised deployment of conversational AI within perioperative care pathways while preserving clinician oversight for verification and patient-specific decision-making.

## INTRODUCTION

Informed consent is a foundational ethical requirement of modern medicine, yet patients frequently undergo surgery without a functional understanding of anaesthesia risk^1,2^. This failure is not unique to any single healthcare system: across settings, patient comprehension of anaesthesia risk is shaped by barriers of language, literacy, and time, and its role in clinical decision-making remains rarely measured and even more rarely addressed. The gap between the recognised importance of patient-reported understanding and the absence of scalable solutions to achieve it has persisted for decades, and defines the central problem this study addresses.

Large-scale studies consistently demonstrate poor retention of perioperative information, with comprehension further diminished by age, language barriers, and low health literacy^3–5^. The consequences are not trivial: inadequate understanding is associated with heightened anxiety, impaired decision-making capacity, and reduced satisfaction with care^6,7^. The central failure of contemporary consent is therefore not the absence of information, but the failure of information transfer^8^. Despite decades of recognition, the architecture of preoperative education has changed little. Information is still delivered synchronously by clinicians during time-limited encounters conducted under competing clinical pressures, leaving patients a limited opportunity to process, question, or consolidate complex risk information.

Adjunctive approaches, including written materials, instructional videos, and nurse-led education, improve knowledge retention modestly, but their effects remain incremental rather than transformative^9,10^. Critically, none alter the underlying structure of information delivery: education remains clinician-initiated, encounter-dependent, and constrained by the cognitive and temporal limitations of a single clinical interaction^11^. As surgical volumes increase and perioperative services face escalating workforce pressures, this model becomes progressively more difficult to sustain.

Digital health interventions have begun to redistribute elements of perioperative care beyond the physical clinic^12^. Virtual assessment pathways achieve outcomes and patient satisfaction comparable to face-to-face consultation^13^, demonstrating that aspects of perioperative workflow can be redesigned without compromising safety. However, these systems largely preserve the assumption that clinicians remain the primary agents of patient education. Conversational AI introduces a fundamentally different model. Unlike static digital tools, conversational systems can engage patients in iterative, self-paced dialogue, adapt explanations dynamically to individual comprehension, and deliver structured, retrieval-grounded information across multiple languages and at scale^14^. In this framework, patient-reported understanding is no longer constrained to the clinical encounter, but can instead be established upstream of clinician contact.

Interest in clinical applications of large language models (LLMs) has expanded rapidly^15,16^, with early evidence demonstrating substantial utility in clinician-facing domains, including documentation^17^, decision support^18^, and anaesthesia risk stratification^19^. In contrast, evidence for patient-facing deployment remains limited and conceptually underdeveloped^20^. Existing randomised trials of chatbot-based preoperative interventions have focused primarily on anxiety reduction^21,22^, rather than whether AI systems can reliably establish understanding of medical risk. Moreover, these systems largely relied on general-purpose conversational models without retrieval-augmented grounding in institutional consent frameworks, limiting both clinical specificity and reliability.

Recent evidence further suggests that model capability alone is insufficient to ensure safe or effective patient outcomes. In a recent randomised study, lay users assisted by general-purpose LLMs performed no better than controls, and in some tasks performed worse, despite the same models demonstrating high diagnostic accuracy when queried directly by investigators^23^. Failure arose not from deficiencies in medical knowledge, but from breakdowns at the human-AI interface, including incomplete information provision by users, ineffective communication of AI-generated advice, and failure of patients to appropriately contextualise model outputs. These findings suggest that the principal challenge in patient-facing AI is no longer whether LLMs possess medical knowledge, but whether clinical systems can reliably translate that knowledge into patient-reported understanding and safe action.

We evaluated this question through a randomised controlled trial of PEAR (Preoperative Education of Anaesthesia Risks), a multilingual retrieval-augmented conversational AI system grounded in institutional consent materials. We hypothesised that PEAR would achieve equivalent patient-reported understanding of anaesthesia risk compared with standard consultation while substantially improving the efficiency and scalability of preoperative education. More fundamentally, we sought to determine whether meaningful patient comprehension could be established before clinician contact, and whether this redefines the role of the clinical consultation itself: from a primary site of information delivery to a secondary layer of verification, contextualisation, and shared decision-making within perioperative care.

## RESULTS

### Participants and enrolment

Between January and April 2026, a total of 130 patients were recruited, with 65 in each arm. In the intervention group, the mean age was 49.9 ± 12.6 years; 55.4% were male. American Society of Anesthesiologists (ASA) Physical Status Classification System was ASA 1 in 4.6%, ASA 2 in 75.4%, and ASA 3 in 20.0%. The education level was primary or secondary in 17.0%, pre-university or above in 83.1%. The majority of patients (84.6%) completed PEAR in English; 13.8% used the Chinese-language interface; and 1.5% used Malay. No patient chose Tamil interaction. Patients self-completed the PEAR in 96.9% of sessions; two elderly patients had their questionnaire completed by a caregiver. Baseline characteristics are summarized in Table 1.

**Table 1.** Baseline characteristics.

| Characteristic | Intervention (n = 65) | Control (n = 65) | p-value |
| --- | --- | --- | --- |
| Age, years (mean ± SD) | 49.9 ± 12.6 | 54.8 ± 16.1 | 0.052 |
| Male sex, n (%) | 36 (55.4) | 35 (53.8) | 0.873 |
| **ASA physical status, n (%)** |  |  | 0.692 |
| ASA 1 | 3 (4.6) | 2 (3.1) |  |
| ASA 2 | 49 (75.4) | 53 (81.5) |  |
| ASA 3 | 13 (20.0) | 10 (15.4) |  |
| **Highest education, n (%)** |  |  | 0.904 |
| Primary | 4 (6.2) | 5 (7.7) |  |
| Secondary | 7 (10.8) | 8 (12.3) |  |
| Pre-university | 15 (23.1) | 14 (21.5) |  |
| University | 39 (60.0) | 38 (58.5) |  |
| **Primary language, n (%)** |  |  | 0.642 |
| English | 55 (84.6) | 50 (76.9) |  |
| Chinese | 9 (13.8) | 13 (20.0) |  |
| Others | 1 (1.5) | 2 (3.1) |  |
| **User, n (%)** |  |  | - |
| Self-completed | 63 (96.9) | - |  |
| Caregiver-completed | 2 (3.1) | - |  |
| **Consulted by, n (%)** |  |  | >0.999 |
| Doctor | 49 (75.4) | 48 (73.8) |  |
| Nurse | 16 (24.6) | 17 (26.2) |  |
| **Surgery type, n (%)** |  |  | 0.988 |
| Orthopaedics | 25 (38.5) | 27 (41.5) |  |
| General surgery | 17 (26.2) | 16 (24.6) |  |
| Urology | 7 (10.8) | 6 (9.2) |  |
| Gynaecology | 7 (10.8) | 6 (9.2) |  |
| Others | 9 (13.8) | 10 (15.4) |  |

SD, standard deviation; ASA, American Society of Anesthesiologists physical status.

### Patient-reported understanding after anaesthesiologist consultation

Post-consultation patient-reported understanding met the pre-specified equivalence criterion on all three primary items (Figure 1). The mean between-group difference (PEAR − control) was −0.03 (90% CI −0.19 to +0.13) for Item 1, −0.05 (90% CI −0.22 to +0.13) for Item 2, and +0.12 (90% CI −0.07 to +0.32) for Item 3. For all comparisons, the 90% confidence intervals lay entirely within the pre-specified equivalence margin of ±0.5 Likert points (all TOST p < 0.001, Bonferroni-adjusted α = 0.0167). In supportive analyses, all lower bounds of the corresponding 95% confidence intervals remained above the pre-specified non-inferiority margin of −0.5 (all one-sided p < 0.001). Two-sided Mann–Whitney U tests, reported for completeness, were non-significant on all items (p = 0.85, 0.89, 0.40 for Items 1, 2, and 3, respectively) (Supplementary Table 2). The composite score (mean of the three items) yielded a between-group difference of +0.02 (90% CI −0.14 to +0.18), also satisfying the pre-specified equivalence criterion (TOST p < 0.001); internal consistency was high (Cronbach’s α = 0.87).

**Figure 1.**
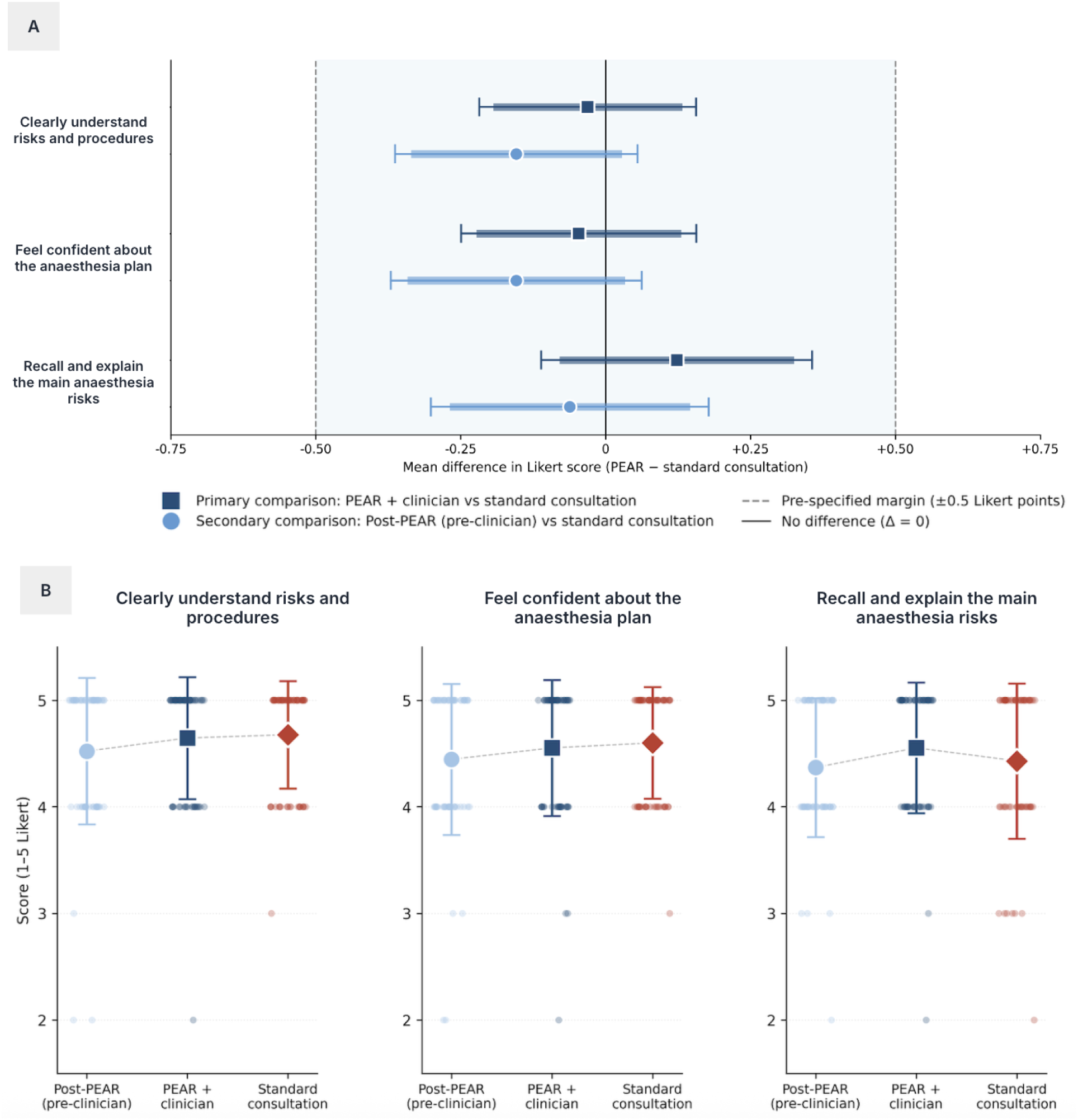
Patient-reported understanding of anaesthetic risk after PEAR-augmented versus standard preoperative consultation. (A) Forest plot of between-group mean differences (PEAR − standard consultation) for the three primary 5-point Likert items (n = 65 per arm). The shaded region indicates the pre-specified equivalence margin (Δ = ±0.5 Likert points). Dark blue squares represent the primary comparison (PEAR + clinician consultation versus standard consultation); light blue circles represent the pre-specified secondary comparison (post-PEAR scores before clinician consultation versus standard consultation). (B) Distribution of patient-reported understanding scores across study groups and assessment time points. Points represent individual patient responses; larger markers indicate group means with ±1 SD.

### Pre-consultation educational effect of PEAR

The intervention group used PEAR for a mean of 16.6 ± 7.2 minutes (median 15 minutes, range 5–35) before their scheduled consultation. Post-PEAR understanding scores (assessed using the same three primary outcome items described above, each on a 5-point Likert scale) were 4.52 ± 0.69, 4.45 ± 0.71, and 4.37 ± 0.65 for Items 1, 2, and 3, respectively, captured before any clinician contact. Comparing these scores directly against control group post-consultation scores showed no statistically significant difference across all three items (p > 0.05), indicating that PEAR alone achieved patient-reported understanding scores comparable to those observed after standard face-to-face consultation. (Supplementary Table 2).

Within the intervention group, the subsequent clinician consultation conferred a small but statistically significant further improvement in Item 1 (clearly understand risks and procedures; p = 0.046) and Item 3 (can recall and explain main risks; p = 0.007), while Item 2 (feel confident about the anaesthesia plan) did not reach significance (p = 0.108). These findings are consistent with the known value of direct question-and-answer interaction for consolidating recall of specific risk information, while confirming that PEAR established a strong educational foundation independently of clinician contact.

### Technology acceptance

Technology acceptance was high across all four TAM constructs, with all composite scores exceeding the 3.5/5 adoption threshold^24^ (Table 2).

**Table 2.**
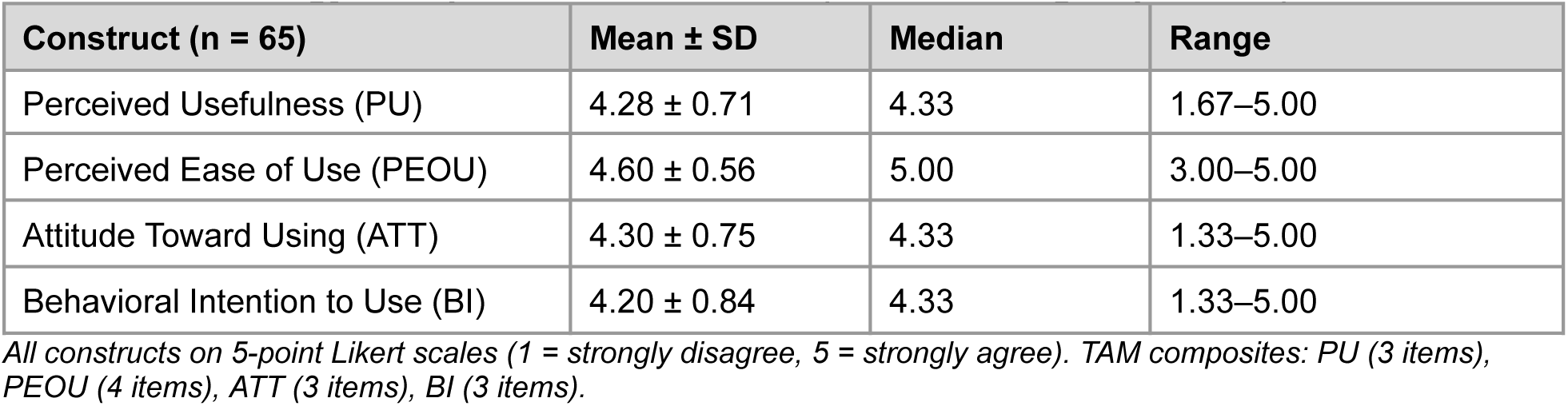
Technology Acceptance Model scores (intervention group, n = 65)

| Construct (n = 65) | Mean ± SD | Median | Range |
| --- | --- | --- | --- |
| Perceived Usefulness (PU) | 4.28 ± 0.71 | 4.33 | 1.67–5.00 |
| Perceived Ease of Use (PEOU) | 4.60 ± 0.56 | 5.00 | 3.00–5.00 |
| Attitude Toward Using (ATT) | 4.30 ± 0.75 | 4.33 | 1.33–5.00 |
| Behavioral Intention to Use (BI) | 4.20 ± 0.84 | 4.33 | 1.33–5.00 |

*All constructs on 5-point Likert scales (1 = strongly disagree, 5 = strongly agree). TAM composites: PU (3 items), PEOU (4 items), ATT (3 items), BI (3 items).*

### Patient and physician preference

Of the 65 intervention-arm participants, 41 (63.1%) preferred PEAR over physician consultation as their primary source of preoperative information. Patients preferring PEAR were younger (47.4 ± 13.3 versus 54.2 ± 10.5 years; p = 0.038), had lower ASA physical status scores (2.02 versus 2.39; p = 0.003), and were more commonly English-speaking, suggesting that younger and healthier patients were more inclined to engage autonomously with AI-delivered education. On Technology Acceptance Model (TAM) assessment, PEAR-preferring participants reported significantly greater ease of use, more favourable attitudes toward the system, and stronger behavioural intention to use AI-assisted education tools, whereas perceived usefulness did not differ significantly between groups, indicating that usability rather than perceived utility was the principal driver of adoption (Figure 2).

**Figure 2.**
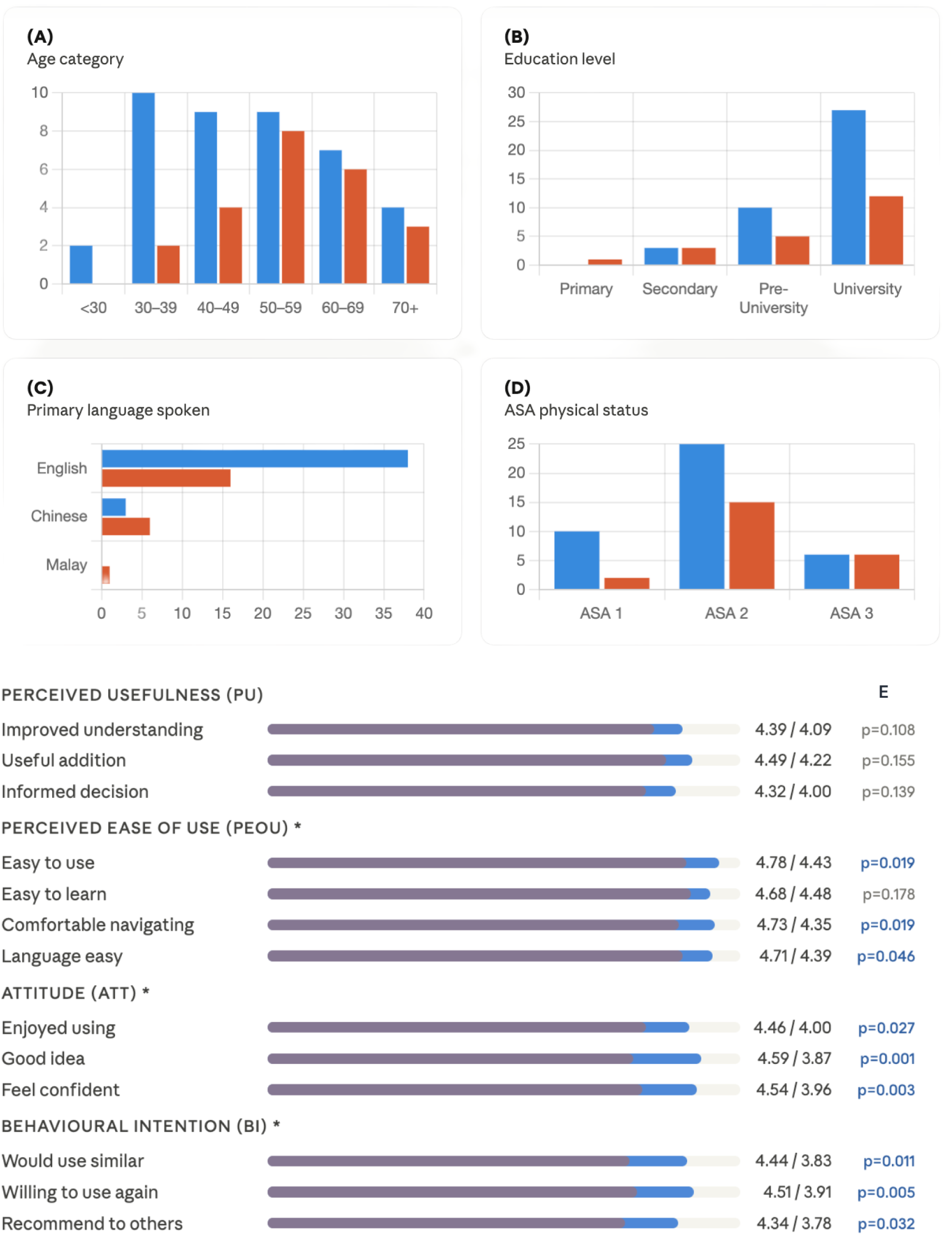
Patient characteristics and Technology Acceptance Model (TAM) scores stratified by information delivery preference. (A-D) Demographic and clinical characteristics of intervention-arm participants, grouped by preferred information delivery method: (A) age category, (B) education level, (C) primary language spoken, and (D) ASA physical status classification. Blue represents participants preferring PEAR; Red represents those preferring in-person physician consultation. (E) TAM domain scores (1-5 Likert scale) comparing preference groups. Scores are displayed as layered horizontal bars: the purple segment indicates the mean score for physician consultation preference, and the blue segment is overlaid to show the mean score for PEAR preference. Because PEAR scores were uniformly higher across all items, the visible blue extension beyond the purple bar visually represents the absolute difference in scores favoring PEAR. Values are reported as means (PEAR/physician consultation).

The most frequently discussed topics during PEAR interactions related to fasting instructions, pain management, and postoperative recovery concerns (Supplementary Table 3 and 4). Qualitative feedback was generally positive, although some participants expressed concerns regarding accuracy, liability, and the need for continued human oversight. Documentation inaccuracies or clinically inappropriate recommendations were identified in 11 of 65 sessions (16.9%); all were classified as minor or moderate in severity, with no major or catastrophic events observed (Supplementary Table 7).

### Clinical quality

PEAR demonstrated clinically meaningful pattern recognition. This included unprompted identification of symptoms suggestive of obstructive sleep apnoea in three patients who had not raised this concern, detection of recent upper respiratory tract infection contraindicating elective surgery, and flagging of a drug allergy disclosed in the questionnaire but absent from the electronic health record. PEAR identified and escalated 40 of 42 retrospectively adjudicated clinically significant findings present within the conversational transcripts.

### Clinical time efficiency

Implementation of PEAR reduced documentation time per encounter to 9.25 ± 4.90 min (95% CI: 8.04–10.46) versus a historical institutional benchmark of 19.35 ± 10.95 min18, representing a mean saving of 10.10 min per encounter (52.2% reduction; 95% CI: 7.13–13.07). Total patient contact time decreased from 21.31 ± 10.22 min to 12.12 ± 6.52 min (95% CI: 10.50–13.74; Δ = −9.19 min; 43.1% reduction; 95% CI: 6.19–12.19). Combined, PEAR reduced total clinical time per patient by 19.29 min (95% CI: 15.06–23.52).

### Conversational quality and text readability

Across all PEAR transcripts, mean readability corresponded to grade level 14.9 ± 2.1 across four validated indices, substantially above the recommended 6th–8th grade target for patient-facing educational materials25. Despite this, patients achieved high self-reported understanding scores, suggesting that interactive conversational delivery may partially mitigate limitations associated with lexical complexity in static written materials. However, adaptive tailoring of educational complexity to patient literacy or educational level was not implemented in the current system and remains an important direction for future development. Patient Education Materials Assessment Tool for Printable Materials (PEMAT-P) assessment demonstrated high transcript understandability (98.8 ± 4.3%), although actionability scores were lower (56.5 ± 7.6%). Additional conversational metrics and detailed scoring are provided in the Supplementary Table 5.

### Economic analysis

All costs are presented in 2026 Singapore dollars (SGD) with US dollar (USD) equivalents converted at an exchange rate of 1 SGD = 0.79 USD (as of May 2026). Applying a weighted clinician compensation rate of SGD 125.24 h⁻¹ (USD 98.94; derived from 75.4% physician time at SGD 140 h⁻¹ and 24.6% nursing time at SGD 80 h⁻¹), the observed time savings corresponded to SGD 40.26 (USD 31.81) per encounter (95% CI: SGD 31.44–49.09). At an estimated annual volume of 25,000 encounters at Singapore General Hospital (SGH), a 1,800-bed tertiary academic medical centre, this equates to gross annual savings of SGD 1.01 million (USD 0.80 million; 95% CI: SGD 0.79–1.23 million), representing 6,060 physician-hours and 1,977 nursing-hours reallocated from documentation to direct care. Assuming a standard full-time equivalent (FTE) workload of 2,000 h yr⁻¹^26^, these efficiencies correspond to an approximate reduction of 3.0 physician FTEs and 1.0 nursing FTE. Annual service provision costs totalled SGD 15,600 (USD 12,324), comprising SGD 600 for cloud infrastructure hosting, SGD 5,000 for variable interaction-based computing costs at SGD 0.20 per encounter, and SGD 10,000 for systems integration and maintenance^27^, corresponding to an estimated net annual operational benefit of approximately SGD 0.99 million (USD 0.78 million) and a return on investment exceeding 6,300% (Table 3). Sensitivity analyses confirmed robustness of these estimates across plausible variations in encounter volume, clinician compensation, and efficiency parameters (Supplementary Table 6).

**Table 3.**
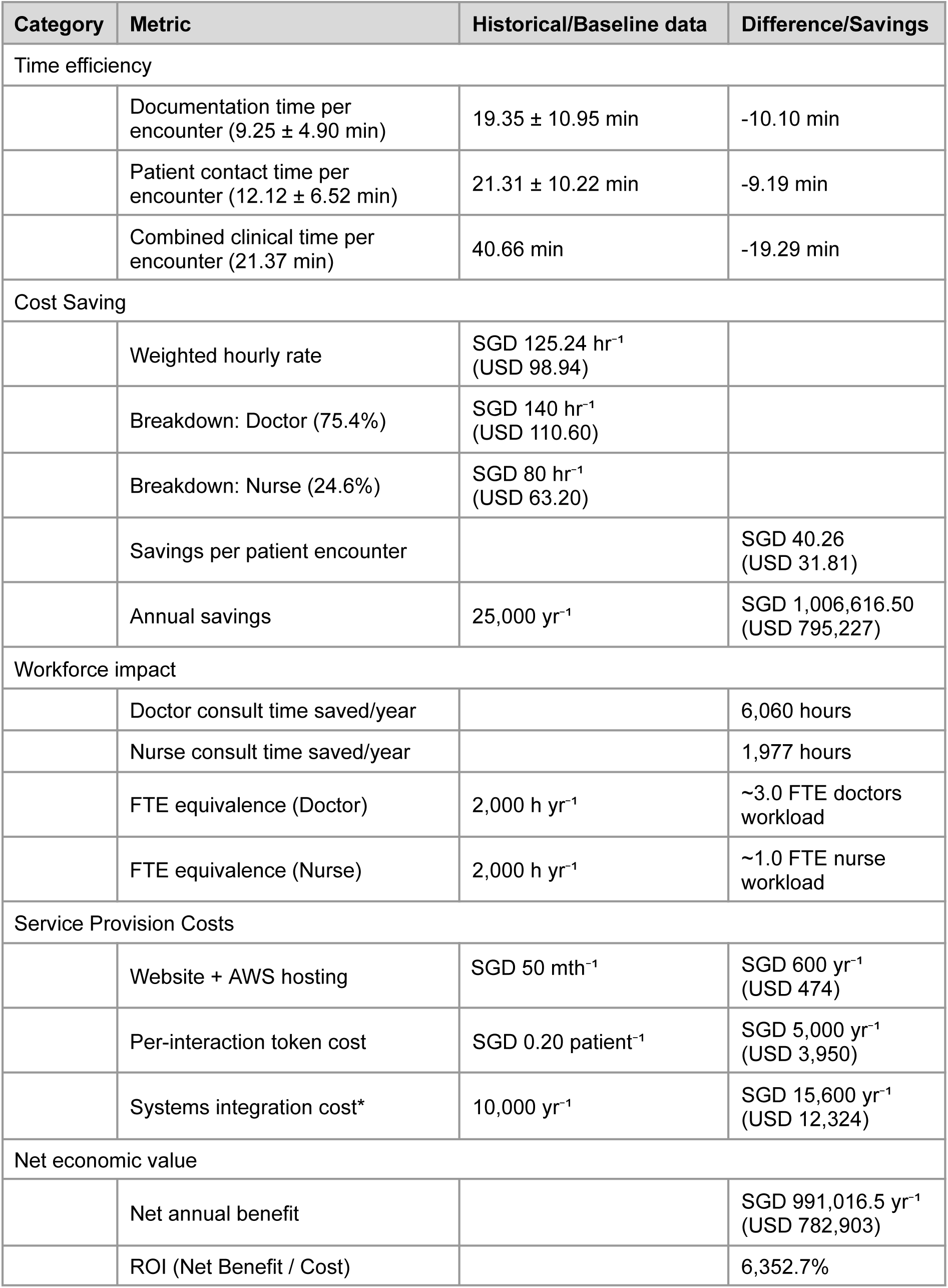
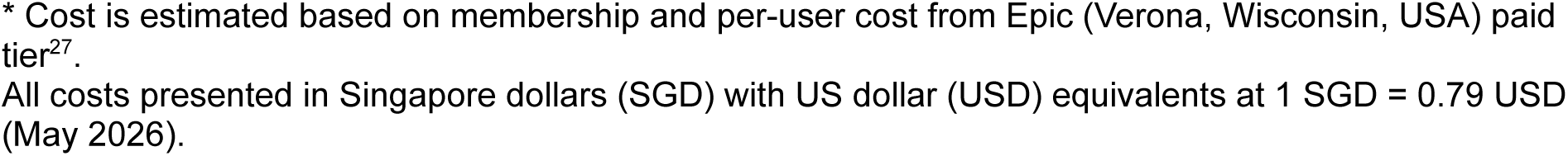
Economic analysis of PEAR-augmented clinical care.

| Category | Metric | Historical/Baseline data | Difference/Savings |
| --- | --- | --- | --- |
| Time efficiency |  |  |  |
|  | Documentation time per encounter (9.25 ± 4.90 min) | 19.35 ± 10.95 min | -10.10 min |
|  | Patient contact time per encounter (12.12 ± 6.52 min) | 21.31 ± 10.22 min | -9.19 min |
|  | Combined clinical time per encounter (21.37 min) | 40.66 min | -19.29 min |
| Cost Saving |  |  |  |
|  | Weighted hourly rate | SGD 125.24 hr<sup>-1</sup><br>(USD 98.94) |  |
|  | Breakdown: Doctor (75.4%) | SGD 140 hr<sup>-1</sup><br>(USD 110.60) |  |
|  | Breakdown: Nurse (24.6%) | SGD 80 hr<sup>-1</sup><br>(USD 63.20) |  |
|  | Savings per patient encounter |  | SGD 40.26<br>(USD 31.81) |
|  | Annual savings | 25,000 yr<sup>-1</sup> | SGD 1,006,616.50<br>(USD 795,227) |
| Workforce impact |  |  |  |
|  | Doctor consult time saved/year |  | 6,060 hours |
|  | Nurse consult time saved/year |  | 1,977 hours |
|  | FTE equivalence (Doctor) | 2,000 h yr<sup>-1</sup> | ~3.0 FTE doctors workload |
|  | FTE equivalence (Nurse) | 2,000 h yr<sup>-1</sup> | ~1.0 FTE nurse workload |
| Service Provision Costs |  |  |  |
|  | Website + AWS hosting | SGD 50 mth<sup>-1</sup> | SGD 600 yr<sup>-1</sup><br>(USD 474) |
|  | Per-interaction token cost | SGD 0.20 patient<sup>-1</sup> | SGD 5,000 yr<sup>-1</sup><br>(USD 3,950) |
|  | Systems integration cost* | 10,000 yr<sup>-1</sup> | SGD 15,600 yr<sup>-1</sup><br>(USD 12,324) |
| Net economic value |  |  |  |
|  | Net annual benefit |  | SGD 991,016.5 yr<sup>-1</sup><br>(USD 782,903) |
|  | ROI (Net Benefit / Cost) |  | 6,352.7% |

\* Cost is estimated based on membership and per-user cost from Epic (Verona, Wisconsin, USA) paid tier<sup>27</sup>. All costs presented in Singapore dollars (SGD) with US dollar (USD) equivalents at 1 SGD = 0.79 USD (May 2026).

### Human oversight: exploratory safety and economic analysis

Although PEAR demonstrated strong educational performance and substantial workflow efficiency gains, an important translational question is whether clinician oversight remains necessary or whether autonomous deployment could be justified on safety and economic grounds. To address this, we performed a prospective error analysis comparing PEAR-generated consultation summaries against independent clinician review of corresponding conversational transcripts.

Across 65 sessions, attending clinicians identified 11 documentation errors (16.9% of encounters), comprising four minor and seven moderate-severity events, with no major or catastrophic errors observed (Supplementary Table 7). Errors included AI-generated hallucinations, clinically inappropriate recommendations, and documentation inaccuracies inconsistent with patient-reported information or institutional guidelines. All identified errors originated from the AI system rather than patient non-disclosure. At the conversational exchange level, 11 erroneous outputs were identified across 670 assistant turns, corresponding to an error frequency of 1.6%.

To explore the implications of autonomous deployment at scale, we extrapolated the observed error rates to an estimated annual workload of 25,000 preoperative encounters. Under these assumptions, PEAR would be expected to generate approximately 4,250 documentation errors annually. Applying severity-stratified cost estimates yielded projected cumulative error-related costs ranging from SGD 723,530 to SGD 1.40 million (USD 571,589 to USD 1.11 million) per year, consuming an estimated 73–141% of projected net time-efficiency savings and thereby rendering fully autonomous deployment economically unfavourable under most plausible scenarios (Table 4). Break-even analyses suggested that sustained cost-beneficial autonomous deployment would likely require error rates below thresholds reliably achievable without clinician verification (Figure 3 and Supplementary Table 6).

**Figure 3:**
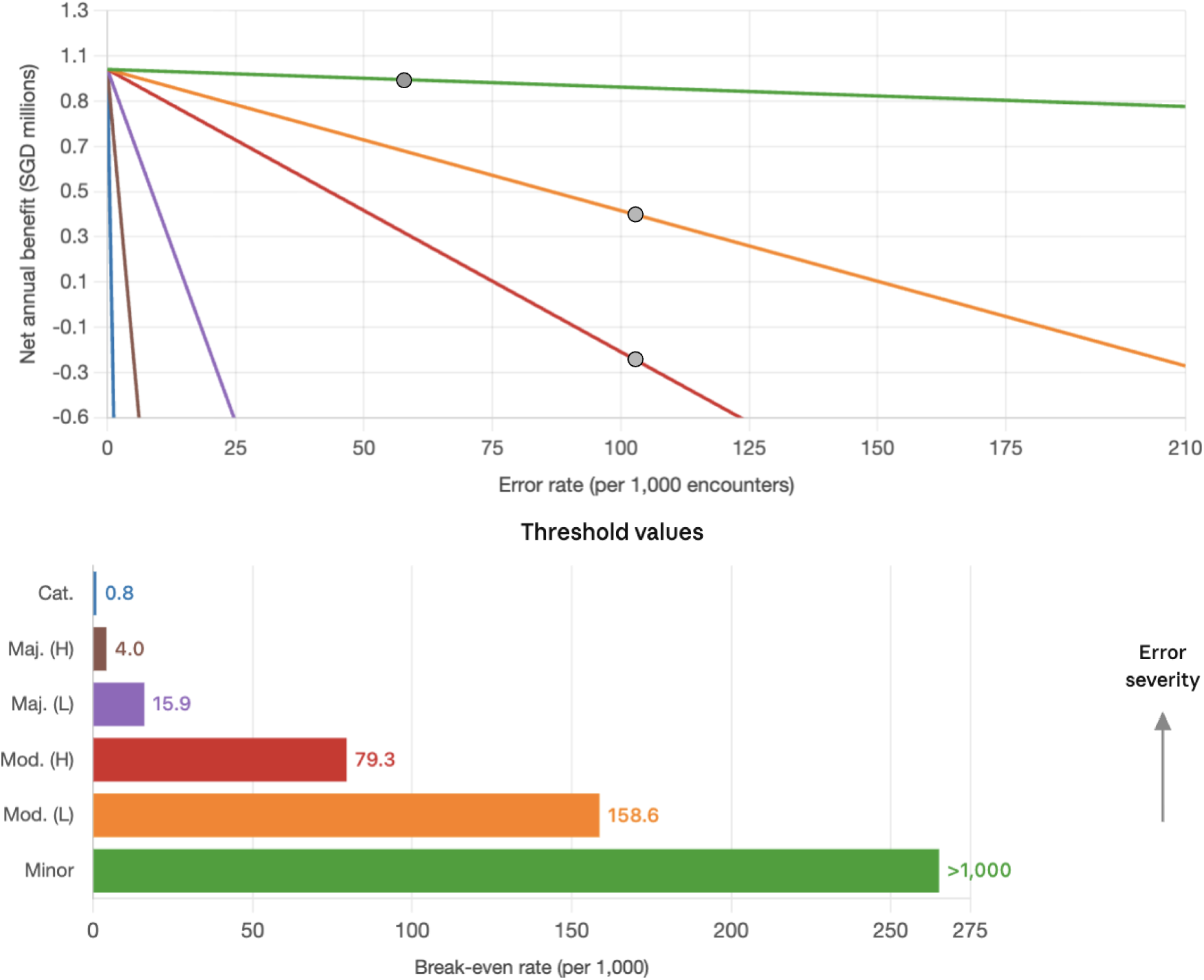
Economic robustness of autonomous PEAR to documentation errors across error severity scenarios. (Upper panel) Net annual benefit (SGD millions) of deploying autonomous PEAR as a function of documentation error rate (per 1,000 encounters), modelled across six error severity scenarios assuming 25,000 annual encounters and a base net saving of SGD 991,016.50 per year. Each line represents a severity category defined by estimated cost per error: Minor (SGD 31.31), Moderate low (SGD 250), Moderate high (SGD 500), Major low (SGD 2,500), Major high (SGD 10,000), and Catastrophic (SGD ≥50,000). The dashed horizontal line denotes the break-even threshold (net benefit = 0), below which error costs exceed PEAR net savings. The grey dots represent the error rates identified in the current retrieval-augmented PEAR system. (Lower panel) Break-even error rates by severity category, calculated as SGD 991,016.50 ÷ (25,000 × cost per error) × 1,000.

**Table 4.** Error severity analysis for autonomous PEAR operation without human oversight.

| Severity | Definition | Observed rate* | Est. Cost per Error (SGD / USD)† | Break-Even Error Rate‡ | Annual Error Cost (SGD)/ USD) | % of Net Savings§ |
| --- | --- | --- | --- | --- | --- | --- |
| Minor | Documentation requires clinician rework (<15 min); no clinical impact | 6.2% | SGD 31.31 / USD 24.74 | 1,266 errors / 1,000 encounters | SGD 48,530 / USD 38,339 | 4.9% |
| Moderate | Delayed care, additional tests, or additional referrals | 10.7% | SGD 250–500 / USD 198–395 | 159-79 errors / 1,000 encounters | SGD 675,000–1.35 million / USD 533,250–1.07 million | 68.1 - 136.2% |
| Major | Adverse clinical event, litigation risk, or significant rework | - | SGD 2,500–10,000 / USD 1,975–7,900 | 16-4 errors / 1,000 encounters | - | - |
| Catastrophic | Severe harm, regulatory penalty, or reputational damage | - | SGD ≥50,000 / USD ≥39,500 | ≤1 error / 1,000 encounters | - | - |
| Total |  | 16.9% |  |  | SGD 723,530–1.4 million / USD 571,589–1.10 million | 73.0 - 141.1% |

All costs are presented in Singapore dollars (SGD) with US dollar (USD) equivalents at 1 SGD = 0.79 USD (May 2026).
\* Observed error rates were derived from independent clinical evaluators reviewing 65 conversational transcripts against corresponding PEAR-generated summaries.
† Cost estimates were derived from three sources: clinician rework time costed at the weighted institutional rate of SGD 125.24 h<sup>-1</sup> (USD 98.94); international proxies for documentation-error-related care delays; and Singapore-adjusted adverse event costs applying healthcare consumer price index and purchasing power parity conversion.
‡ Break-even error rate represents the maximum tolerable error frequency at which cumulative error costs fully offset PEAR's net economic benefit, calculated as: net annual savings (SGD 991,017; USD 782,903) divided by annual encounter volume (25,000) multiplied by cost per error.
§ Percentage of PEAR's net annual savings consumed by projected error-related costs. Values exceeding 100% indicate net economic loss under autonomous operation.
Base parameters: time saved per encounter = 21.37 min (0.356 h); weighted clinician rate = SGD 125.24 h<sup>-1</sup> (USD 98.94); gross savings per encounter = SGD 40.26 (USD 31.81); annual service provision cost = SGD 15,600 (USD 12,324); net annual savings = SGD 991,017 (USD 782,903). Errors are assumed to be independent and additive; costs are not modelled to compound or cascade across encounters.

## DISCUSSION

This randomised trial demonstrates that a retrieval-augmented conversational AI can achieve equivalent patient-reported understanding of anaesthesia risk compared with standard preoperative consultation, while reducing total clinical time per encounter by over 40%. A notable finding from the pre-specified secondary analysis was that patients who interacted with PEAR before clinician contact achieved self-reported understanding scores comparable to those observed after standard clinician consultation alone. This suggests that foundational patient-reported understanding may be established before clinician contact, potentially allowing clinicians to focus consultations on contextualisation, clarification, and shared decision-making.

The marginal benefit conferred by clinician consultation in the intervention arm was largely confined to risk recall rather than conceptual understanding. This pattern is consistent with the known role of direct dialogue in consolidating patient knowledge^28^ and supports a model in which AI delivers standardised baseline education while clinicians focus on contextualisation, uncertainty management, and individual decision-making. Whether this reallocation of informational labour translates into improvements in downstream perioperative outcomes or post-operative patient satisfaction remains to be established.

The projected efficiency gains are meaningful at the institutional scale. Annual savings of approximately SGD 1 million (USD 0.78 million) at a single hospital, with annual service provision costs under SGD 16,000 (USD 12,500), reflect a favourable cost-benefit profile where marginal cost per additional patient interaction approaches zero after initial development^29^. These estimates are broadly consistent with prior evidence from LLM-assisted clinician tools in perioperative medicine^18^ and patient-facing domains^30^. However, the economic model assumes that clinician time freed by PEAR can be redeployed within the existing workforce. In healthcare systems where surgical demand already exceeds workforce capacity, time savings may instead translate into reduced wait times and earlier access to surgery, outcomes with additional clinical and economic value that were not captured in the present analysis. Future evaluations should incorporate demand-side constraints, waitlist costs, and system-level capacity effects to provide a more comprehensive assessment of AI-assisted perioperative workflows.

The value of PEAR extends beyond direct cost savings. Redistribution of routine informational and documentation tasks to an AI layer may reduce administrative burden for clinicians while allowing consultations to focus on contextualisation, uncertainty management, and shared decision-making^31^. From the patient perspective, asynchronous self-paced education may support autonomous decision-making and improve the quality of the consent encounter, although these outcomes require prospective evaluation. Because the underlying retrieval-augmented framework is not specialty-specific, similar models may be applicable to other clinical pathways involving structured education, risk disclosure, or consent processes.

However, the efficiency gains observed in this study remain contingent on continued clinician oversight. Clinicians identified omissions or inaccuracies in 16.9% of encounters, consistent with known limitations of retrieval-augmented systems and patient-facing human–AI interaction workflows^23^. Exploratory modelling suggested that autonomous deployment would substantially erode the projected economic benefit under plausible error scenarios, reinforcing clinician verification as a structural component of safe implementation rather than a temporary safeguard.

Conversational AI systems for patient education introduce important governance and safety considerations^32^. Although PEAR demonstrated a low per-exchange error frequency, clinicians identified omissions or inaccuracies in 16.9% of encounters, consistent with recognised limitations of patient-facing human–AI interaction workflows^20,33^. Accordingly, PEAR was not designed for autonomous deployment. All AI-generated outputs were reviewed and verified by the consulting anaesthesiologist before consent finalisation, with clinician oversight functioning as a structural component of the workflow rather than a temporary safeguard. Safe deployment at scale will require formal governance frameworks incorporating auditability, continuous quality monitoring, version-controlled retrieval systems, and clear clinician accountability for AI-assisted decisions^34,35^. Conversational AI may nevertheless prove particularly valuable in settings with constrained perioperative workforce capacity^36^, where scalable multilingual educational systems could reduce consultation burden and improve access to structured informed-consent processes, although adaptation to local regulatory, linguistic, and cultural contexts would remain necessary.

Patient preference data suggest that implementation may need to be risk-stratified. Preference for AI-mediated education was associated with younger age, lower ASA physical status, and English as a primary language. AI-augmented educational pathways may therefore be most appropriate initially for lower-risk patients, while higher-risk or linguistically diverse patients continue to receive earlier clinician engagement. Importantly, multilingual systems may also reduce comprehension disparities for non-English speakers, provided translation quality and cultural appropriateness are rigorously validated.

### Limitations

Several limitations warrant consideration. This was a single-centre study conducted in a tertiary institution with a predominantly well-educated, English-proficient population, which may limit generalisability to settings with greater linguistic, educational or socioeconomic heterogeneity. The primary endpoint was based on self-reported understanding rather than objective knowledge assessment; patients may overestimate comprehension or retention of complex perioperative information. Accordingly, the extent to which PEAR scores reflect true knowledge acquisition, as opposed to perceived understanding, remains uncertain and should be evaluated in future studies incorporating objective testing. PEMAT-P evaluation was performed only on PEAR-generated educational transcripts, and direct comparison against standard institutional consent materials or clinician-delivered consultation was not conducted. In addition, although educational attainment was recorded at baseline, the study was not powered to determine whether comprehension outcomes differed across education levels or health-literacy strata.

The multilingual analyses were underpowered for definitive subgroup inference, and the applicability of these findings to low- and middle-income settings or health systems with different consent practices requires further validation. In addition, the study was not designed to evaluate downstream perioperative outcomes, including anxiety, decisional conflict, adherence, or postoperative recovery. Time-efficiency comparisons relied on a historical rather than concurrent control, introducing the possibility of temporal confounding from changes in workflow, staffing, or case complexity. The magnitude of the observed efficiency gains should therefore be interpreted as approximate rather than definitive, and prospective studies with concurrent controls are needed to establish the true operational impact of deployment. In addition, the economic analysis did not model demand-side workforce constraints, surgical waitlist effects, or downstream system-level impacts of increased perioperative capacity.

The study also did not evaluate the cognitive demands associated with sustained clinician review of AI-generated outputs. The long-term sustainability of clinician-in-the-loop oversight, including potential vigilance fatigue or over-reliance during large-scale deployment, warrants prospective evaluation in future implementation studies^37^.

### Conclusions

This trial demonstrates that retrieval-augmented conversational AI can support scalable preoperative patient education within a supervised clinical workflow. PEAR established patient-reported understanding comparable to standard consultation while substantially reducing clinician time requirements, supporting a model in which foundational perioperative education may occur before direct clinician contact. These findings provide a reproducible framework for integrating conversational AI into consent-dependent clinical pathways while preserving clinician oversight for verification, contextualisation, and shared decision-making.

## METHODS

### Study design and setting

We conducted a prospective, single-centre, open-label, two-arm randomised controlled trial at the Preoperative Evaluation Clinic of Singapore General Hospital (SGH), a 1,800-bed tertiary referral centre performing approximately 25,000 elective surgical procedures annually. All patients undergoing elective surgery attend preoperative anaesthesia assessment. The study was conducted from January to April 2026. The protocol was approved by the SingHealth Centralised Institutional Review Board (CIRB 2025/0673) and registered at ClinicalTrials.gov (NCT06949462). All participants provided written informed consent. The study is reported in accordance with CONSORT 2025 guidelines^38^. A CONSORT flow diagram is provided in Supplementary Figure 1.

### Participants and Randomisation

Eligible participants were adults (≥21 years) with ASA physical status I - III scheduled for elective low- to moderate-risk surgery, who were able to interact in English, Chinese, Malay, or Tamil. Participants were required to have sufficient cognitive capacity to engage with study procedures as assessed by the consenting clinician. Exclusion criteria included emergency or high-risk surgery, inability to provide informed consent, and active psychiatric or neurological conditions impairing decision-making capacity. Randomisation was conducted using a 1:1 block design using a computer-generated block randomisation sequence with variable block sizes. Blinding of participants and treating clinicians was not feasible due to the nature of the intervention. Independent evaluators assessing transcript quality and PEMAT-P scores were blinded to study allocation and hypotheses.

### Intervention: PEAR system

PEAR (Preoperative Education of Anaesthesia Risks) is a retrieval-augmented conversational AI system built on a large language model (Claude Haiku 4.5, Anthropic) with multilingual embedding capabilities (Qwen 3). The system integrates three components: (1) a clinician interface for procedural input and consent criteria specification, (2) a structured digital pre-operative questionnaire capturing comorbidities, medications, allergies, symptoms, and functional status, and (3) a conversational interface delivering personalised anaesthesia education through a combination of guided inputs and free-text interaction (Figure 4). The retrieval corpus comprised institutional anaesthetic consent forms, departmental clinical guidelines, and curated patient education materials, maintained under version control to ensure alignment with local practice. Patient questionnaire responses were converted into a structured patient-specific medical context, and all AI-generated educational content was grounded in institutional guidelines through retrieval-augmented generation (RAG) using multilingual embeddings. Multilingual educational content was developed using institutional consent materials and iteratively reviewed by bilingual clinicians and native speakers for clinical accuracy, linguistic clarity, and contextual appropriateness across English, Chinese, and Malay interfaces. The system generated a structured clinical summary indexed to the patient queue number, which was transmitted in real-time into the institutionally isolated electronic health record environment for clinician review. Participants in the intervention arm completed PEAR before their scheduled consultation. Both groups subsequently underwent standard in-person anaesthesiologist consultation.

**Figure 4.**
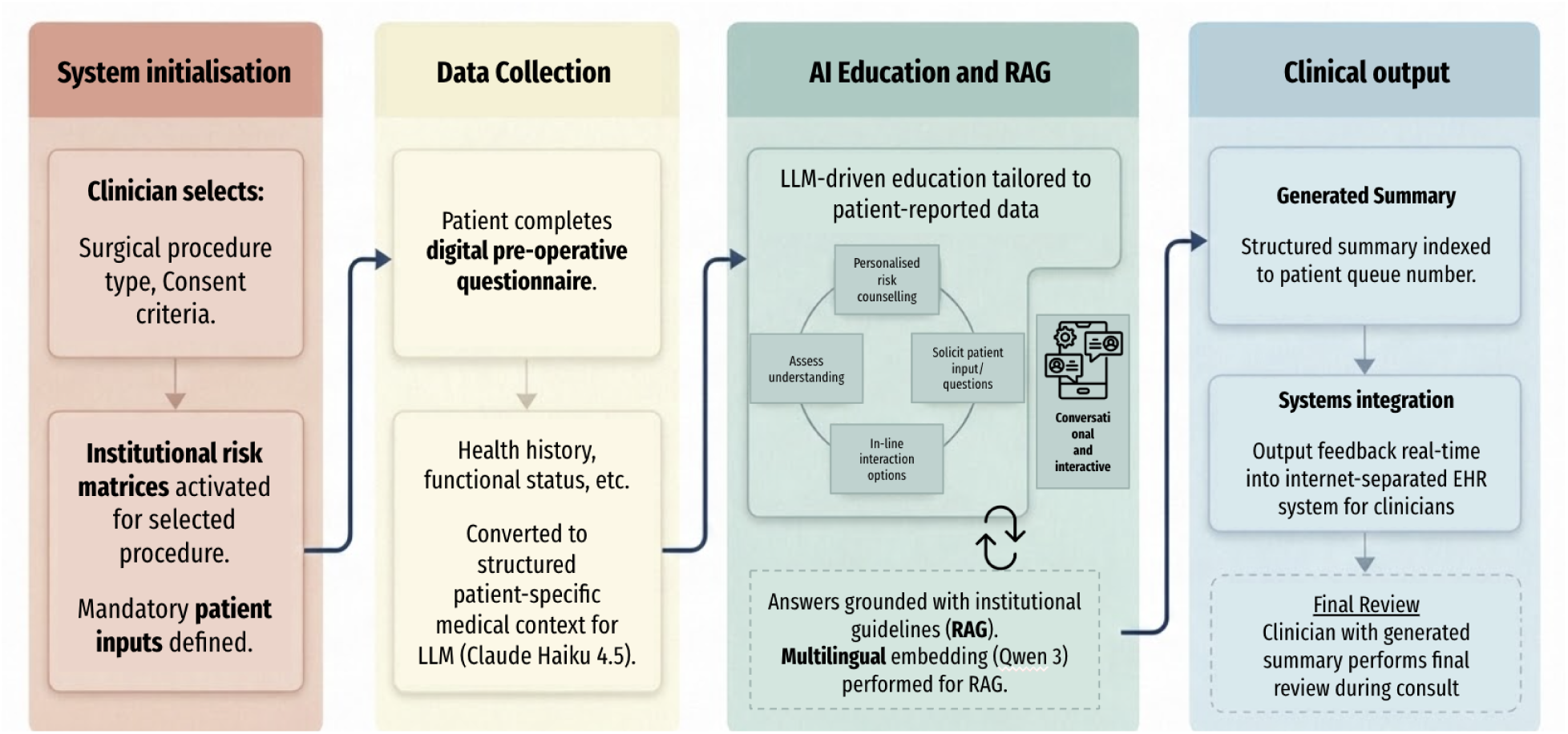
Architecture and workflow of the PEAR (Preoperative Education of Anaesthesia Risks) system. The PEAR workflow comprises four phases: (1) system initialisation, where clinicians define procedure-specific consent requirements; (2) patient data collection through a structured preoperative questionnaire; (3) multilingual retrieval-augmented conversational education using institutional guidelines and patient-specific context; and (4) generation of a structured clinical summary for clinician review within the electronic health record system. All educational outputs were grounded in institutional consent materials through retrieval-augmented generation.

### Outcomes

The primary outcome was patient-reported understanding of anaesthesia risk following the preoperative consultation. This was assessed using three 5-point Likert-scale items evaluating patients’ perceived understanding of anaesthesia risk, confidence in the anaesthesia plan, and ability to recall and explain key risks. In participants randomised to the intervention arm, these measures were additionally collected immediately after completion of the PEAR interaction and before clinician consultation to evaluate pre-consultation educational effects.

Secondary outcomes included multiple domains capturing usability, safety, quality, and efficiency. Technology acceptance was assessed using the Technology Acceptance Model, including perceived usefulness, perceived ease of use, attitude towards use, and behavioural intention. Patient preference for AI-mediated versus in-person education was evaluated using a structured trade-off scenario. Safety and clinical appropriateness of PEAR outputs were assessed in real time by attending clinicians, including identification of omissions, hallucinations, and overall suitability for clinical use. Hallucinations were operationally defined as AI-generated statements unsupported by either patient-provided information or the institutional retrieval corpus. Independent evaluation of transcript quality was performed using the Patient Education Materials Assessment Tool for printable materials (PEMAT-P), with two blinded consultant anaesthesiologists assessing understandability and actionability. Additional transcript-level quality metrics included clinical accuracy, appropriateness of perioperative advice, and identification of clinically relevant findings.

Prospective measurement of consultation and documentation time in the control arm was not pre-specified in the trial protocol, as the efficiency analysis was intended primarily to estimate the magnitude of time savings attributable to PEAR relative to standard unassisted practice. Accordingly, the comparator benchmark was derived from a contemporaneous institutional study conducted in the same preoperative clinic under identical workflow conditions and involving the same cohort of consultant anaesthesiologists^18^, providing the closest available estimate of routine clinical practice. Clinically significant findings were defined as any symptom, medication, allergy, or medical history with the potential to alter anaesthetic management or prompt further perioperative evaluation (e.g., loose tooth requiring further dental review). The presence of such findings was determined through retrospective review of complete conversational transcripts by two blinded consultant anaesthesiologists, who served as the reference standard.

Interaction characteristics were analysed using system-generated logs, including the number of conversational turns, the distribution of input modalities (structured versus free text), and session duration. Text readability of AI-generated responses was assessed using multiple validated indices, including Flesch Reading Ease, Flesch–Kincaid Grade Level, Gunning Fog Index, Coleman–Liau Index, SMOG, and Linsear Write formula. Clinical efficiency outcomes included patient interaction time, clinician consultation time, and total encounter duration. An exploratory economic evaluation was conducted to estimate cost implications at the institutional scale. Error severity was classified using a four-level taxonomy adapted from medical-error frameworks. Minor error refers to documentation inaccuracy requiring clinician rework < 15 minutes, no clinical impact (e.g., misformatted medication entry, missing comorbidity later captured at consultation). Moderate error is defined when prompting additional investigation, referral, or delay in clinical pathway without harm is required (e.g., omitted allergy that altered anaesthetic plan upon clinician verification). Major error refers to those with potential for adverse clinical event, litigation risk, or significant pathway disruption. Catastrophic error are cases which lead to severe patient harm or regulatory breach.

Each conversational transcript and corresponding PEAR-generated summary was independently reviewed by two consultant anaesthesiologists blinded to study allocation and to one another’s ratings. Disagreements were resolved by consensus discussion; a third anaesthesiologist served as adjudicator where consensus could not be reached. Errors were further categorised by mechanism: (i) AI-generated content errors (hallucinations, factually incorrect statements, clinically inappropriate recommendations); (ii) documentation-transfer errors (information present in the conversation but lost, distorted, or mislabelled in the generated summary).

### Statistical analysis

The primary analysis was a pre-specified equivalence comparison of PEAR-augmented consultation versus standard preoperative consultation on each of the three primary patient-reported understanding items. The equivalence margin was set at ±0.5 Likert points, corresponding to the minimally important difference used in the sample-size calculation. Equivalence was assessed using two one-sided tests (TOST) and declared if the 90% confidence interval for the between-group mean difference (PEAR − control) lay entirely within the interval −0.5 to +0.5. As supportive analyses, we additionally evaluated non-inferiority using the same lower margin (−0.5) and performed rank-based comparisons using the two-sided Mann–Whitney U test.

Multiplicity across the three primary items was controlled with a Bonferroni-adjusted α of 0.05/3 = 0.0167. Composite reliability across the three items was assessed using Cronbach’s α; the composite (mean) score was analysed as a pre-specified sensitivity outcome. Within-arm comparisons of pre- and post-clinician scores in the intervention arm used Wilcoxon signed-rank tests. Baseline characteristics were compared using independent t-tests, Mann–Whitney U, or chi-square tests as appropriate. Likert items are presented as both mean ± SD (to permit equivalence inference at the group-mean level) and median (IQR) (to reflect their ordinal nature).

The trial was registered as a comparative randomised controlled trial (NCT06949462) without pre-registration of a specific analytic framework; the non-inferiority and equivalence framework, including the Δ = 0.5 margin, was finalised in the statistical analysis plan prior to data lock, using the same margin justified in the sample-size calculation. The registration has been amended accordingly. Analyses were conducted on complete cases without imputation in Python 3.11.

### Economic analysis

Economic evaluation was conducted from the institutional perspective, with time savings translated into cost savings using a weighted clinician hourly rate derived from local salary benchmarks. Annual projections were based on an estimated volume of 25,000 preoperative consultations.

An exploratory break-even analysis was performed to estimate the threshold at which error-related costs under hypothetical autonomous deployment would offset efficiency gains. Errors were categorised by severity, and associated costs were estimated based on clinician rework time, international health services data, and Singapore-adjusted economic indices. Sensitivity analyses were conducted across key parameters, including consultation volume, clinician cost, efficiency gains, and infrastructure costs, to assess the robustness of the model.

Further details regarding cost derivation, modelling assumptions, and parameter ranges are provided in the Supplementary Methods.

## ACKNOWLEDGEMENTS

The authors thank the nursing and administrative staff of the Singapore General Hospital Preoperative Evaluation Clinic for facilitating patient recruitment and all participants who gave their time to this trial. We acknowledge the two anaesthesiologists who performed independent PEMAT and transcript quality scoring.

## AUTHOR CONTRIBUTIONS

Y.K. conceived the study, designed the trial, developed the PEAR system, supervised data collection, performed statistical and economic analyses, interpreted the data, and drafted the manuscript. C.N. and J.L. contributed to system development, patient recruitment, data acquisition, transcript analysis, and manuscript revision. J.S, H.R.A., and B.L.S. provided clinical oversight and critically revised the manuscript. J.Z.A., H.S.S.H., J.Y.M.T., H.K.T., L.J. and D.S.W.T. contributed to AI system design and methodological oversight. M.E.H.O., N.L., and D.S.W.T. supervised the project and contributed to study’s conceptualisation, methodology, and critical revision of the manuscript. N.L. additionally supervised the statistical methodology and economic modelling. All authors reviewed, edited, and approved the final manuscript.

## ETHICS APPROVAL

SingHealth Centralised Institutional Review Board, CIRB Reference 2025/0673. All participants provided written informed consent before enrolment. The study was conducted in accordance with the Declaration of Helsinki.

## DATA AVAILABILITY

De-identified primary data and statistical analysis code will be made available from the corresponding author on reasonable request following full publication.

## Use of AI writing tool

Large language models were used solely for limited editorial assistance, including grammatical refinement and improvement of sentence clarity. All scientific content, interpretation, and final manuscript preparation were performed and verified by the authors.

