## Supplementary Table for "Randomised Trial of a Multilingual Conversational AI for Preoperative Education"

**Supplementary Table 1.** Step-by-step workflow of the personalized LLM (PEAR) pre-anaesthetic consultation system.

| **Interface** | **Features** |
| --- | --- |
| 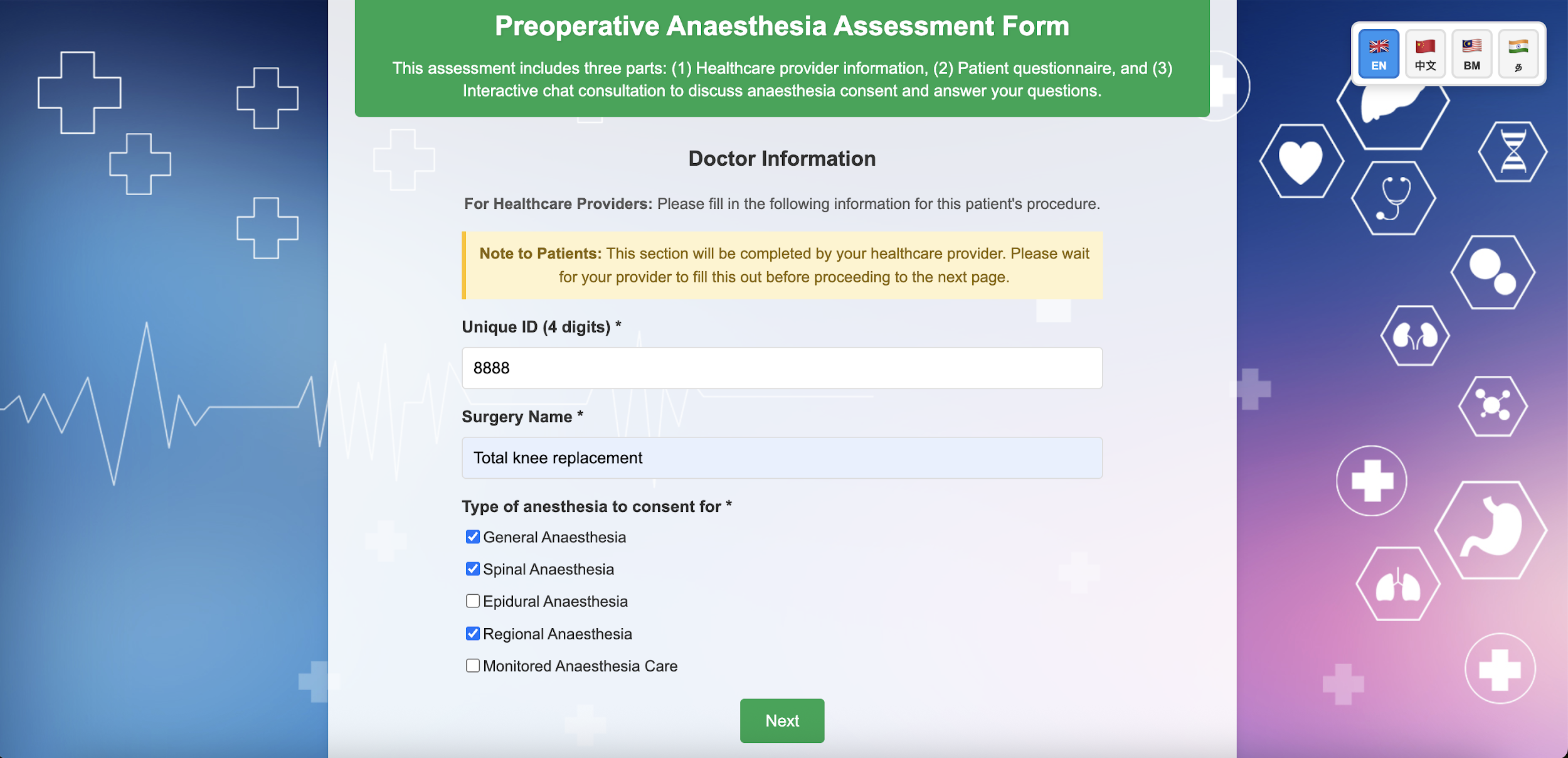 | Clinician inputs patient-specific procedural data and required consent items (e.g., General Anaesthesia, Regional Anaesthesia) to initialize the pre-consultation flow. |
| 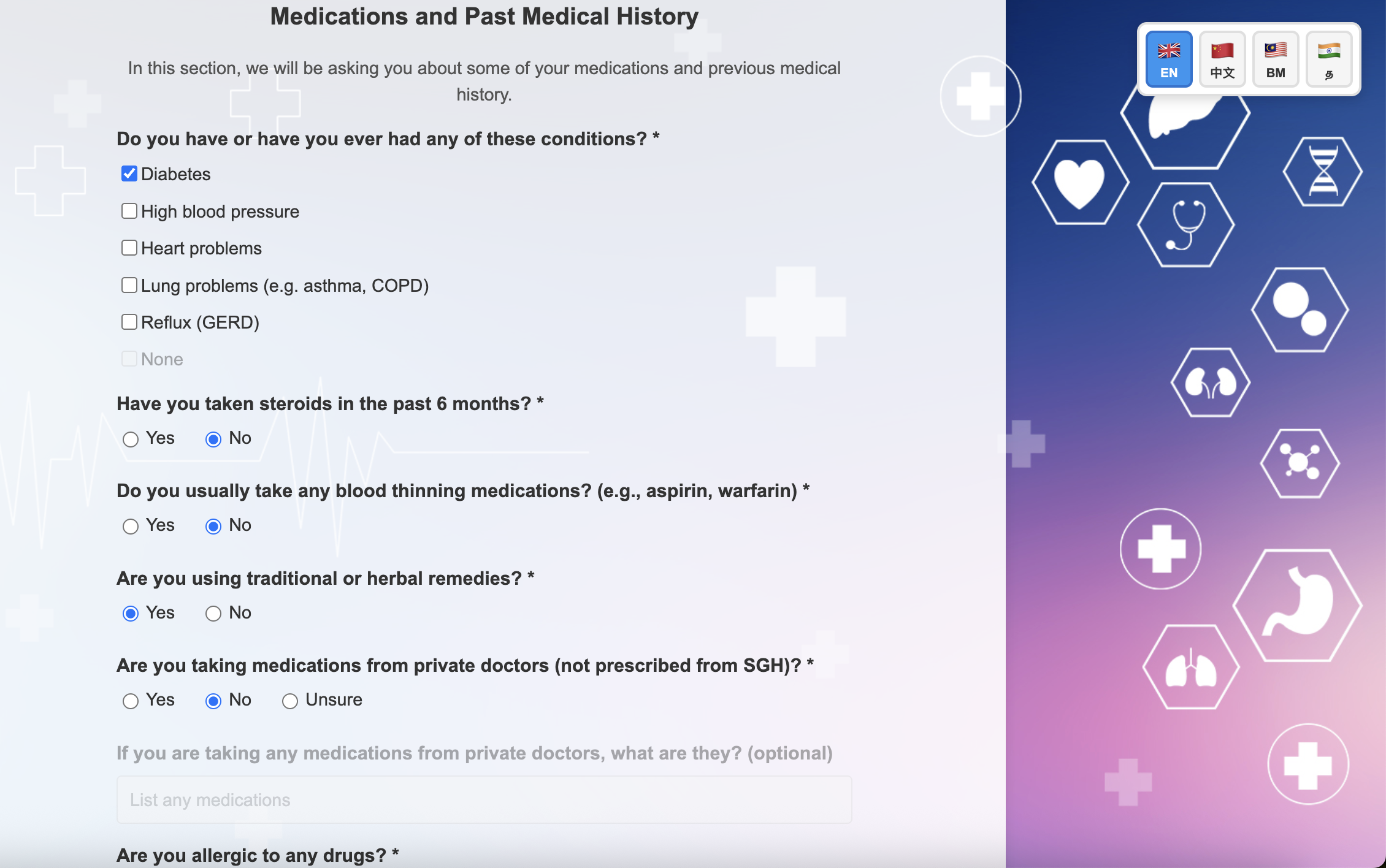 | Patient completes a structured, pre-anaesthetic risk assessment questionnaire, mirroring standard clinical history-taking. |
| 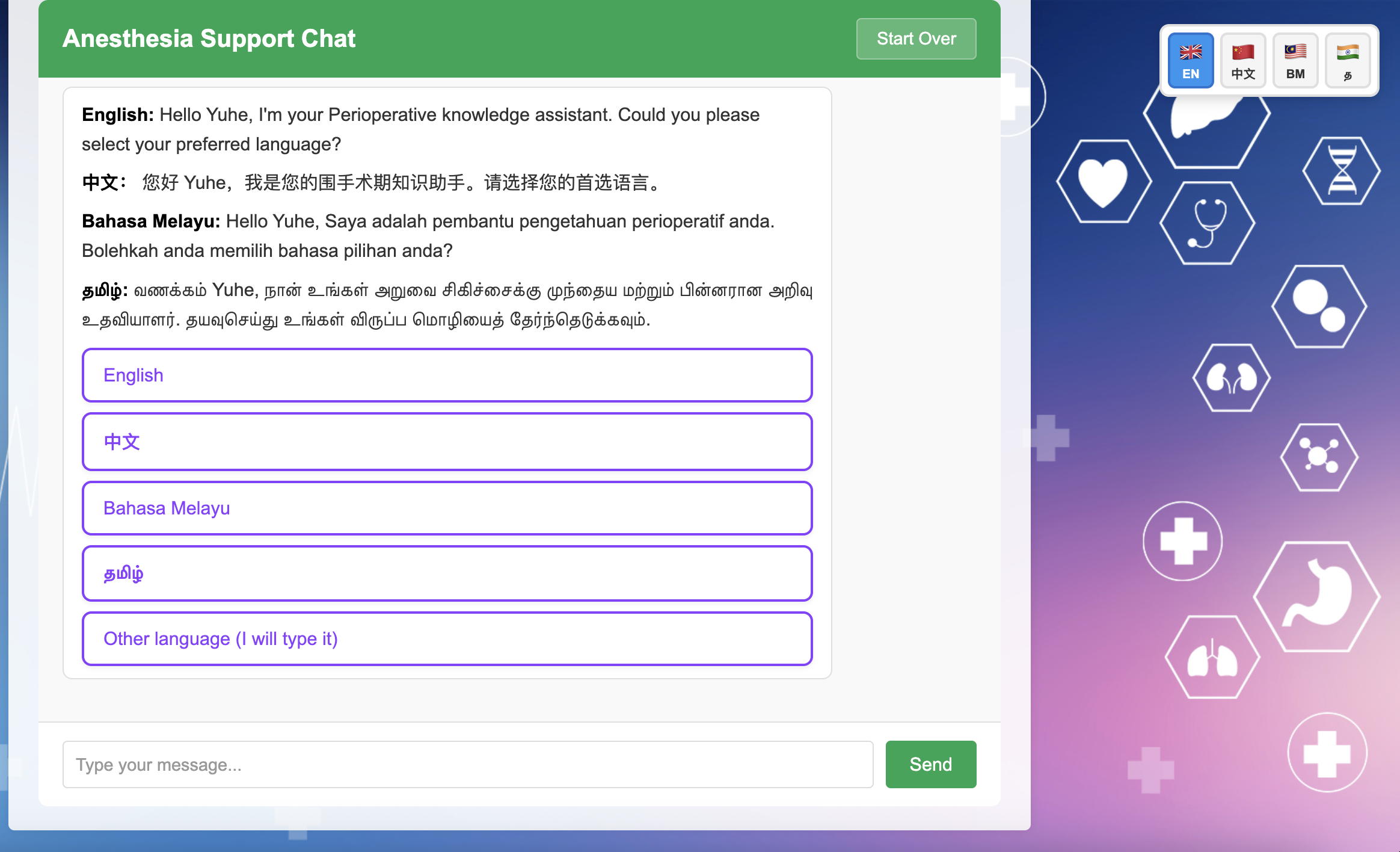 | Post-assessment, the patient initiates an interactive session with the personalized, multilingual Large Language Model (PEAR) |
| 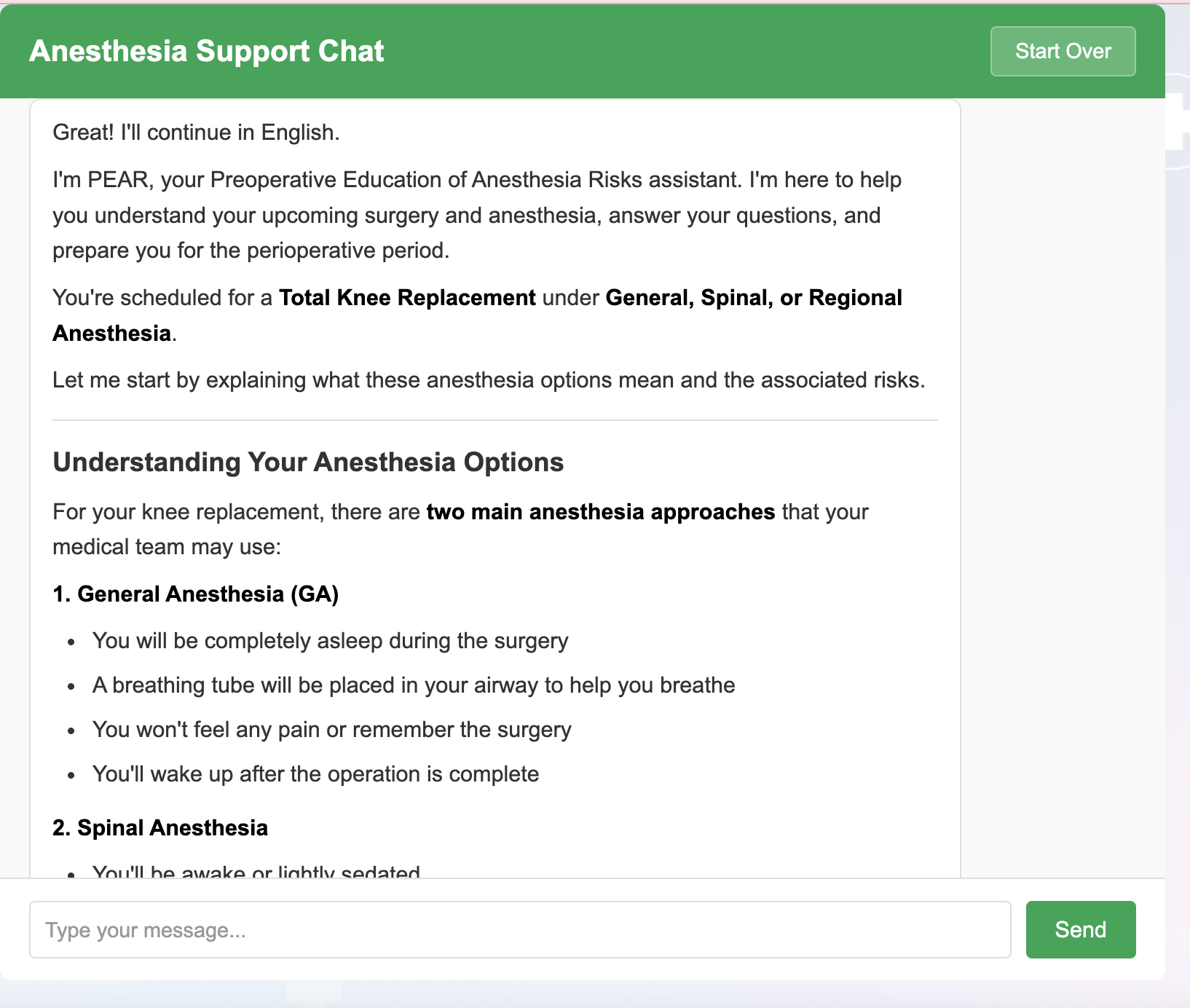 | PEAR automatically initiates the consultation by presenting relevant anaesthesia options tailored to the planned surgical procedure. |
| 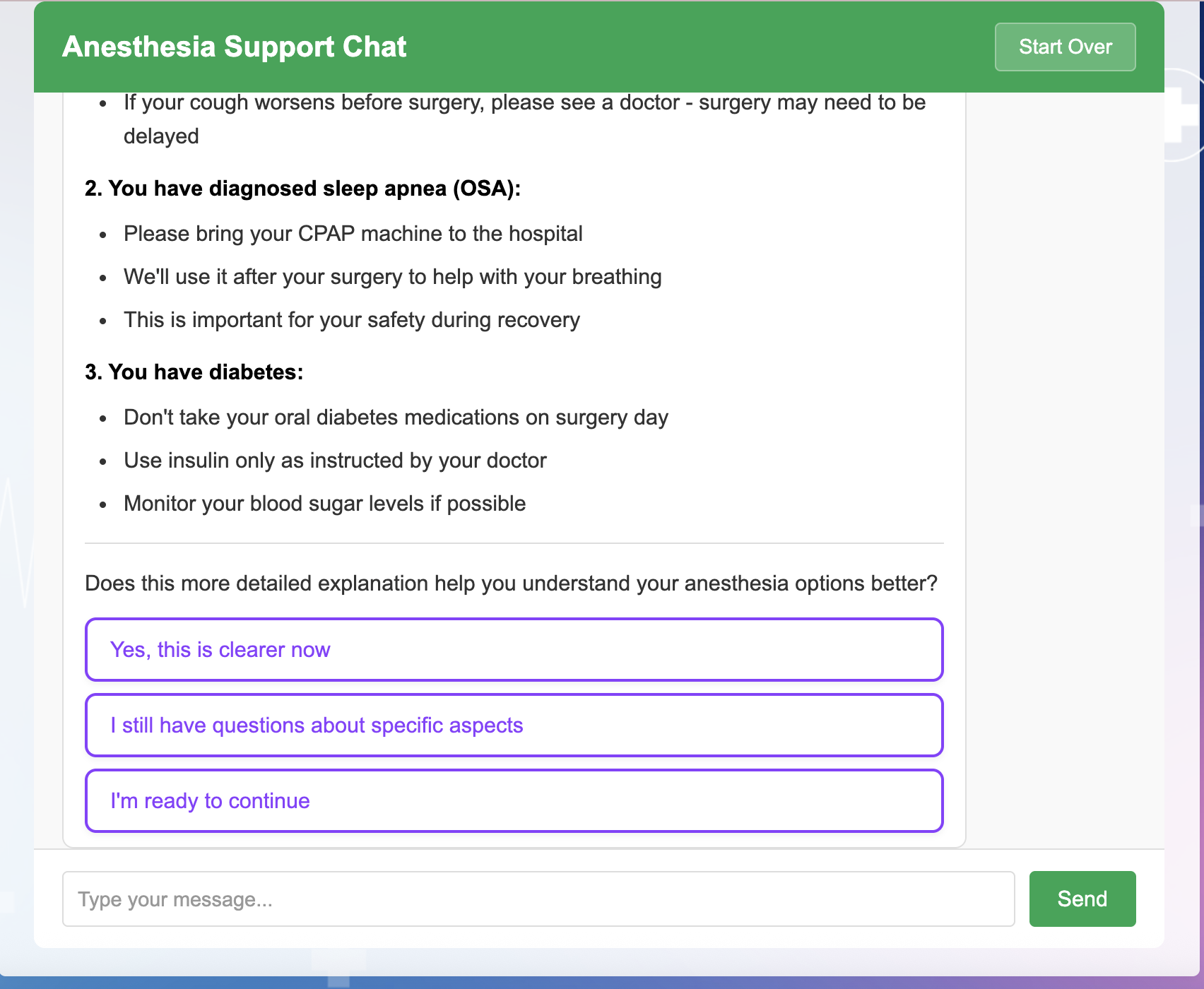 | The LLM delivers personalized risk communication (e.g., addressing anaesthesia risks associated with comorbidities like sleep apnea or diabetes mellitus) via free-text input or structured, inline prompt selections. |
| 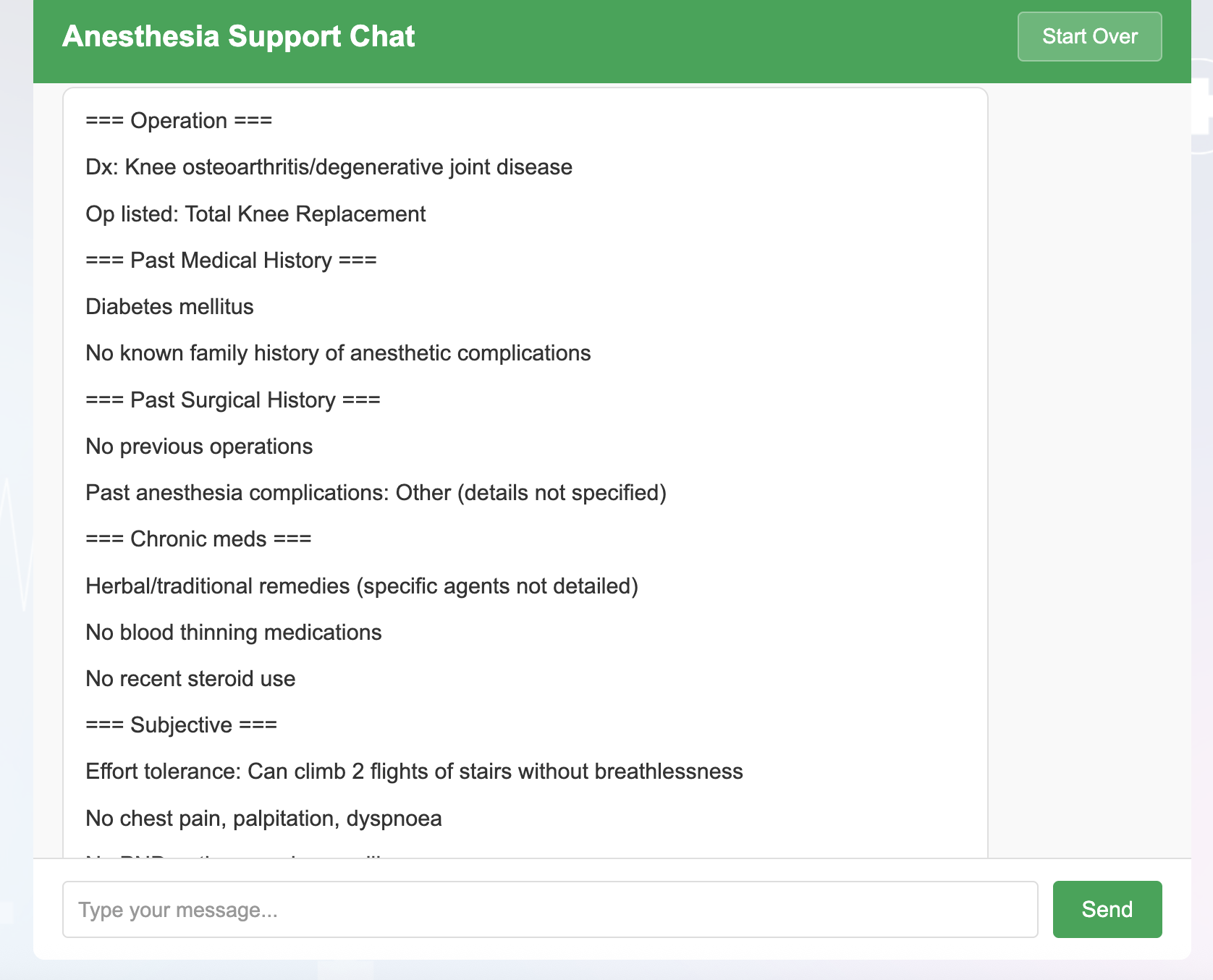 | A structured, free-text clinical summary is automatically generated in a format compatible with physician documentation, and instantly transmitted to the attending physician in real-time, allowing direct copy-pasting into electronic healthcare record system. |

**Supplementary Table 2.** Primary outcome: patient-reported understanding of anaesthetic risk after preoperative consultation, by study arm.

| **Item** | **PEAR***ᵃ* | **Control***ᵃ* | **Diff (95% CI)***ᵇ* | **NI p-valueᶜ** | **TOST p-valueᵈ** | **MW p-value***ᵉ* |
| --- | --- | --- | --- | --- | --- | --- |
| 1: Clearly understand | 4.65 ± 0.57; median 5 (IQR 4–5) | 4.68 ± 0.50; median 5 (IQR 4–5) | −0.031 (−0.218 to +0.156) | <0.001 | <0.001 | 0.85 |
| 2: Feel confident | 4.55 ± 0.64; median 5 (IQR 4–5) | 4.60 ± 0.52; median 5 (IQR 4–5) | −0.046 (−0.249 to +0.157) | <0.001 | <0.001 | 0.89 |
| 3: Recall and explain | 4.55 ± 0.61; median 5 (IQR 4–5) | 4.43 ± 0.73; median 5 (IQR 4–5) | +0.123 (−0.111 to +0.357) | <0.001 | <0.001 | 0.40 |
| Composite*ᶠ* | 4.59 ± 0.57 | 4.57 ± 0.50 | +0.015 (−0.170 to +0.201) | <0.001 | <0.001 | — |

*ᵃ Values are mean ± SD and median (interquartile range) for each Likert item; n = 65 per arm.*

*ᵇ Mean difference (PEAR − control) with two-sided 95% confidence interval, computed by Welch's two-sample t-test.*

*ᶜ One-sided p-value for the non-inferiority test (H₀: mean difference ≤ −0.5; H₁: mean difference > −0.5). Bonferroni-adjusted significance threshold across the three primary items: α = 0.0167.*

*ᵈ Two one-sided test (TOST) p-value for equivalence against the symmetric margin of ±0.5 Likert points; reported as max(p_lower, p_upper).*

*ᵉ Two-sided Mann–Whitney U test, reported as a supportive rank-based comparison; not interpreted as evidence of equivalence or non-inferiority.*

*ᶠ Composite score is the per-patient mean of the three primary items, analysed as a pre-specified sensitivity outcome. Internal consistency across the three items (Cronbach's α) = 0.87 in the pooled sample (n = 130).*

**Supplementary Table 3:** Patient inquiry topics during PEAR interaction.

| **Topic Category** | **Specific Subtopics Included** | **Frequency** |
| --- | --- | --- |
| **Pain Management** | Post-op pain expectations, analgesia options, opioid concerns | 35 (53.8%) |
| **Fasting Guidelines** | Pre-op fasting duration, clear fluids, medication timing | 29 (44.6%) |
| **Waking Up / Recovery** | What to expect after anaesthesia, timeline, discharge readiness | 28 (43.1%) |
| **General Side Effects** | GA side effects, sore throat, dizziness, fatigue | 23 (35.4%) |
| **Nausea & Vomiting** | Prevention, management, risk factors | 17 (26.2%) |
| **GA-Specific Concerns** | Memory loss, awareness under anaesthesia, dental risks | 9 (13.8)% |
| **OSA / Breathing** | Sleep apnoea risks, breathing precautions, CPAP use | 6 (9.2%) |
| **Pre-op Preparation** | Medications to continue/hold, diet, hygiene, arrival instructions | 5 (7.7%) |
| **Allergies / Medical History** | Latex allergy, drug reactions, comorbidity considerations | 5 (7.7%) |
| **Logistics / Discharge** | Length of stay, overnight admission, post-op diet, follow-up | 5 (7.7%) |
| **Other / Free-Text** | Breastfeeding post-LSCS, neck mobility, URTI symptoms, vitamin use | 28 (43.1%) |

**Supplementary Table 4:** Patient qualitative feedback post-PEAR interaction.

| **Positive Feedback** | **Concerns / Areas for Improvement** |
| --- | --- |
| *Usability & Design*  *• Description: Praise for interface simplicity, font size, ease of navigation*  *• Examples: "Easy to use", "simple to understand", "font size good for all ages"*  *• Frequency: 18 (27.7%)* | *Preference for Human Interaction*  *• Description: Desire for clinician involvement; chatbot seen as supplement, not replacement*  *• Examples: "Prefer human interaction", "best as an addition rather than replacement", "doctor is still needed"*  *• Frequency: 12 (18.5%)* |
| *Information Quality*  *• Description: Appreciation for clarity, detail, or usefulness of content*  *• Examples: "Good information", "detailed and easy to understand", "helped with understanding"*  *• Frequency: 16 (24.6%)* | *Accuracy & Liability*  *• Description: Uncertainty about information reliability or accountability for errors*  *• Examples: "Not sure if it is accurate", "who will be responsible?", "unsure how competent AI is"*  *• Frequency: 6 (9.2%)* |
| *Flexibility*  *• Description: Value placed on free-text option or ability to ask open questions*  *• Examples: "We can ask what we want", "free text option is good when question is not listed"*  *• Frequency: 5 (7.7%)* | *Accessibility Barriers*  *• Description: Challenges for elderly, dyslexic, or low-literacy users; suggestion for audio support*  *• Examples: "Patient is elderly, cannot read", "helpful to have an audio option", "Answers are technical and can be made simpler"*  *• Frequency: 6 (9.2%)* |
| *Institutional Trust*  *• Description: Confidence derived from hospital verification/endorsement*  *• Examples: "Feels nice that it is verified by hospital", "trust chatbot more than internet"*  *• Frequency: 3 (4.6%)* | *Lack of Personalisation*  *• Description: Perception that chatbot lacks empathy, initiative, or tailored engagement*  *• Examples: "Chatbot seems to lack personal touch", "doctor will take initiative to ask questions"*  *• Frequency: ~5 (7.7%)* |
|  | *Usability Friction*  *• Description: Need for assistance to complete forms or navigate the tool*  *• Examples: "Patient needed a lot of help to go through the forms and chatbot"*  *• Frequency: 2 (3.1%)* |

**Supplementary Table 5. Conversation dynamics, text readability, and PEMAT assessment**

| **Variable** | **Mean ± SD** | **Median** | **Range** |
| --- | --- | --- | --- |
| **A. Conversation dynamics (n = 65 sessions)** | | | |
| PEAR interaction time (minutes) | 16.6 ± 7.2 | 15 | 5–35 |
| Assistant turns per session | 10.3 ± 5.5 | 10 | 3–30 |
| Inline (button) selections, % | 88.0% | — | — |
| Free-text (typed) input, % | 12.0% | — | — |
| **B. Text readability — pure assistant text (options excluded)** | | | |
| Flesch Reading Ease (target ≥60) | 36.7 ± 8.5 | 39.0 | −2.4 to 54.1 |
| Flesch–Kincaid Grade Level (target 6–8) | 13.3 ± 2.0 | 12.5 | 9.3–18.0 |
| Gunning FOG Index | 16.3 ± 2.1 | 15.7 | 11.9–21.5 |
| Coleman–Liau Index | 14.9 ± 4.8 | 13.3 | 10.8–31.0 |
| SMOG Index | 15.3 ± 1.5 | 15.0 | 12.4–18.1 |
| Mean grade level (target 6–8) | 14.9 ± 2.1 | — | 11.2–22.1 |
| **C. PEMAT content quality assessment** | | | |
| Understandability, pooled (benchmark ≥70%) | 98.8% ± 4.3% | — | — |
| Actionability, pooled (benchmark ≥70%) | 56.5% ± 7.6% | — | — |
| Inter-rater agreement: mean absolute difference | U: 1.6% A: 4.0% | — | — |

**Supplementary Table 6. Sensitivity cost analysis**

| Scenario / Parameter | Variation Level | Gross Savings (SGD) | Total Service Cost (SGD)† | Net Benefit (SGD) | ROI (%) |
| --- | --- | --- | --- | --- | --- |
| BASE CASE | | | | | |
| Base assumptions | 25,000 encounters; SGD 125.24 hr⁻¹; 19.29 min saved; SGD 0.20 patient⁻¹ | 1,006,616.50 | 15,600 | 991,016.50 | 6,353% |
| A. Annual Encounter Volume | | | | | |
| Encounter volume | 15,000 (−40%) | 603,900.00 | 13,600 | 590,300.00 | 4,340% |
|  | 20,000 (−20%) | 805,200.00 | 14,600 | 790,600.00 | 5,415% |
|  | 25,000 (Base) | 1,006,616.50 | 15,600 | 991,016.50 | 6,353% |
|  | 30,000 (+20%) | 1,207,800.00 | 16,600 | 1,191,200.00 | 7,176% |
|  | 35,000 (+40%) | 1,409,100.00 | 17,600 | 1,391,500.00 | 7,906% |
| B. Weighted Clinician Hourly Rate | | | | | |
| Hourly rate (SGD) | 100.19 (−20%) | 805,250.00 | 15,600 | 789,650.00 | 5,062% |
|  | 112.72 (−10%) | 905,900.00 | 15,600 | 890,300.00 | 5,707% |
|  | 125.24 (Base) | 1,006,616.50 | 15,600 | 991,016.50 | 6,353% |
|  | 137.76 (+10%) | 1,107,250.00 | 15,600 | 1,091,650.00 | 6,998% |
|  | 150.29 (+20%) | 1,208,000.00 | 15,600 | 1,192,400.00 | 7,644% |
| C. Time Savings Efficiency | | | | | |
| Efficiency vs. base | 50% (9.65 min saved) | 503,250.00 | 15,600 | 487,650.00 | 3,126% |
|  | 75% (14.47 min saved) | 755,000.00 | 15,600 | 739,400.00 | 4,740% |
|  | 100% (19.29 min saved) | 1,006,616.50 | 15,600 | 991,016.50 | 6,353% |
|  | 125% (24.11 min saved) | 1,258,250.00 | 15,600 | 1,242,650.00 | 7,966% |
|  | 150% (28.94 min saved) | 1,509,750.00 | 15,600 | 1,494,150.00 | 9,578% |
| D. Per-Interaction Infrastructure Cost | | | | | |
| Cost per patient (SGD) | 0.10 | 1,006,616.50 | 13,100 | 993,516.50 | 7,584% |
|  | 0.20 (Base) | 1,006,616.50 | 15,600 | 991,016.50 | 6,353% |
|  | 0.30 | 1,006,616.50 | 18,100 | 988,516.50 | 5,462% |
|  | 0.50 | 1,006,616.50 | 23,100 | 983,516.50 | 4,258% |
|  | 1.00 | 1,006,616.50 | 35,600 | 971,016.50 | 2,728% |

**Supplementary Table 7.** Characterization of errors identified in PEAR chatbot conversations, stratified by economic severity and potential clinical impact.

| **No.** | **Economic Severity** | **Cost per Error (SGD)†** | **(Language) Error Description** | **Type of error** |
| --- | --- | --- | --- | --- |
| 1 | Minor | ~31.31 | (English) Instructed patient to "bring CPAP machine," when the patient never indicated CPAP use | AI-generated content errors (hallucinations, factually incorrect statements, clinically inappropriate recommendations). |
| 2 | Minor | ~31.31 | (Chinese) Stated tramadol would be avoided post-op to prevent PONV without a clinical indication |  |
| 3 | Minor | ~31.31 | (English) Specified "no milk/dairy after 12 mn" rather than standard "nil by mouth." |  |
| 4 | Moderate | 250–500 | (English) Listed arcoxia/omeprazole as "chronic medications" when discussed only as hypothetical options for pain management |  |
| 5 | Moderate | 250–500 | (English) Recommended dental referral solely based on mention of crowns/bridges |  |
| 6 | Moderate | 250–500 | (English) Suggested booking a post-op ICU bed for an arm flap operation without a clinical indication |  |
| 7 | Minor | ~31.31 | (English)Repetitive prompting for past surgical history after the patient provided the surgery name | Documentation-transfer errors (information present in the conversation but lost, distorted, or mislabelled in the generated summary). |
| 8 | Moderate | 250–500 | (English) Did not probe further when patient reported limited effort tolerance ("walk indoors only") |  |
| 9 | Moderate | 250–500 | (Chinese) Documented "No OSA symptoms" despite patient reporting daytime somnolence; OSA noted only in anaesthesia considerations section |  |
| 10 | Moderate | 250–500 | (English) Stated fasting instructions were "explained" when only summarised in the output |  |
| 11 | Major | 2,500–10,000 | (English) Documented "No known drug allergies" when the patient indicated an allergy, but did not specify the medication |  |
