## Supplementary Methods for "Randomised Trial of a Multilingual Conversational AI for Preoperative Education"

### **Economic evaluation framework**

Economic evaluation was conducted from the institutional perspective to estimate the financial impact of PEAR implementation relative to standard preoperative care. The primary outcome of the economic analysis was net annual cost savings derived from reductions in clinician time, with secondary analyses evaluating the impact of documentation errors under hypothetical autonomous deployment.

### ***Cost parameter derivation***

### ***Clinician time cost***

A weighted clinician hourly rate was derived based on the observed distribution of consultation providers in the study cohort, comprising 75.4% physician-led and 24.6% nurse-led consultations. Unit costs were obtained from Singapore Ministry of Health salary benchmarks, with physician time valued at SGD 140 h⁻¹ and nursing time at SGD 80 h⁻¹. This yielded a weighted average rate of SGD 125.24 h⁻¹, which was applied to time savings to estimate cost reduction per encounter.

***Operating theatre cost (contextual parameter)***

Operating theatre (OT) costs were included as contextual parameters for modelling downstream implications of delays and cancellations. In the absence of Singapore-specific public data, a base estimate of USD 46.04 min⁻¹ was obtained from a published international systematic review^1^. This value was converted to Singapore dollars using World Bank purchasing power parity (PPP) conversion factors and adjusted to 2025 values using the Singapore healthcare consumer price index, yielding an estimated cost of approximately SGD 62 min⁻¹.

***Surgical cancellation cost***

The base cost of a cancelled operation was derived from a European multicentre study reporting EUR 2,400 per cancelled case^2^. This value was converted to Singapore dollars using PPP-adjusted exchange rates and inflated to 2025 values. To reflect the higher resource utilisation associated with same-day cancellations, a multiplier of 2–3× was applied based on prior health services research^3,4^, yielding an estimated range of SGD 5,000 per cancellation for modelling purposes.

***PEAR implementation costs***

Annual system costs were modelled as the sum of fixed and variable components. Fixed costs included cloud infrastructure hosting (SGD 600 yr⁻¹) and system integration and maintenance (SGD 10,000 yr⁻¹)^5^. Variable costs were based on interaction-level compute usage, estimated at SGD 0.20 per encounter, corresponding to SGD 5,000 annually at an institutional volume of 25,000 encounters. Total annual operating cost was therefore estimated at SGD 15,600.

***Time–cost conversion***

Per-encounter cost savings were calculated by multiplying the observed reduction in clinician time (in hours) by the weighted hourly rate. Annual cost savings were derived by scaling per-encounter savings to the institutional case volume of 25,000 consultations. Workforce impact was estimated by converting total time saved into full-time equivalent (FTE) units assuming 2,000 working hours per FTE per year.

***Break-even error threshold analysis***

To evaluate the economic implications of autonomous deployment, we modelled the impact of documentation and clinical errors in the absence of clinician oversight. Errors were stratified into four categories: minor (requiring clinician rework without clinical impact), moderate (resulting in delayed care or additional investigations), major (associated with adverse clinical events), and catastrophic (resulting in severe harm or system-level consequences).

Costs for minor errors were calculated based on clinician rework time. Costs for moderate and major errors were estimated using international health services data, adjusted to the Singapore context using PPP and healthcare inflation indices. Catastrophic error costs were modelled using upper-bound estimates from healthcare liability literature.

The break-even error rate (ε*) was defined as the maximum tolerable error frequency at which cumulative error-related costs equal the net annual savings from time efficiency. This was calculated as:

The break-even error rate (ε*) was calculated as:

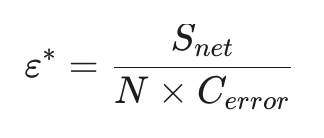

Where:

- *Snet* = Net annual savings from time efficiency (SGD 991,016.50)
- *N* = Annual encounter volume (25,000)
- Cerror = Estimated cost per error (SGD)

This formulation allows estimation of error tolerance thresholds across different severity levels.

***Sensitivity analysis***

Deterministic one-way sensitivity analyses were performed to assess the robustness of economic estimates. Key parameters varied included annual encounter volume (±40%), clinician hourly cost (±20%), magnitude of time savings (50–150% of observed values), and per-interaction computational cost (SGD 0.10–1.00).

For the error-threshold analysis, cost estimates for moderate and major errors were varied by ±50% to account for uncertainty in international proxy data and local cost adjustments.

***Model assumptions***

The economic model was based on several simplifying assumptions. Errors were assumed to occur independently, with additive costs that do not compound or cascade. Time savings and error costs were assumed to scale linearly with encounter volume. Historical institutional benchmarks for consultation time were assumed to be representative of standard care workflows. PPP-adjusted international cost estimates were assumed to reasonably approximate Singapore-specific costs. A one-year time horizon was adopted, and no discounting was applied. Economic break-even thresholds were interpreted independently from clinical safety thresholds, which may be more stringent.

5.	Home - Vendor Services. https://vendorservices.epic.com/.
