## Supplementary figures and images for "Randomised Trial of a Multilingual Conversational AI for Preoperative Education"

### Supplementary Figure

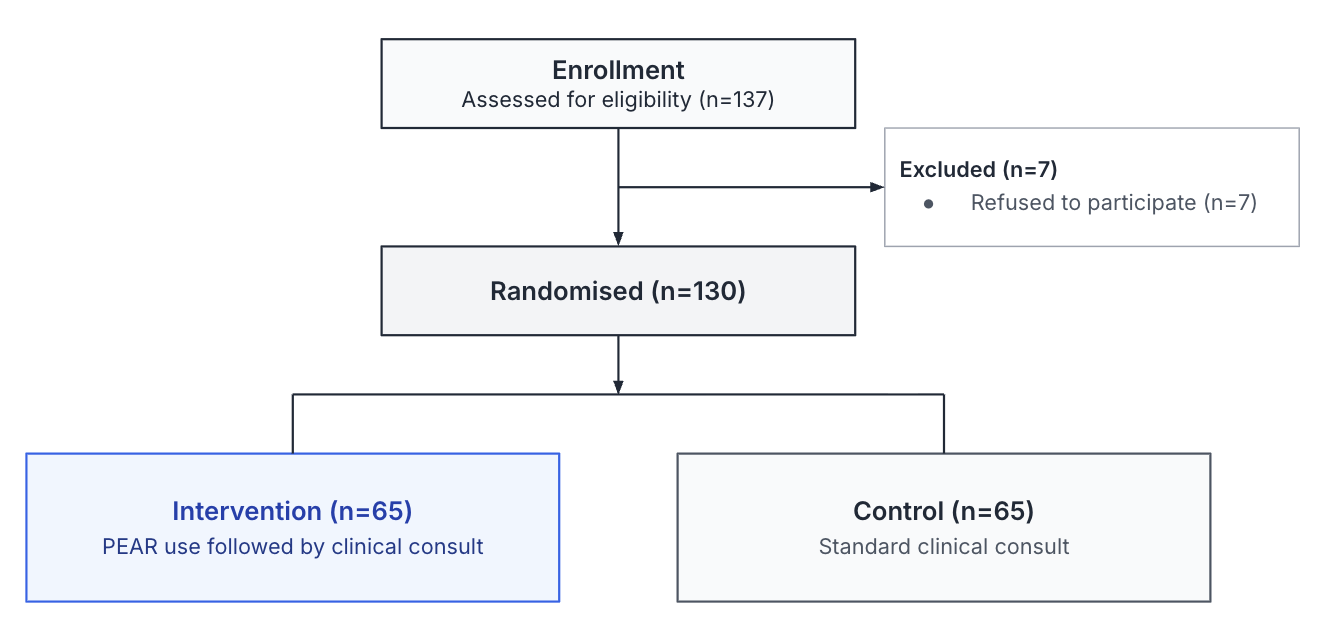

**Supplementary Figure A1.** CONSORT flow diagram.
